# Derivation and Validation of a Machine Learning Approach to Detect and Mitigate Biases in Healthcare Data

**DOI:** 10.1101/2023.11.06.23298164

**Authors:** Faris F. Gulamali, Ashwin S. Sawant, Lora Liharska, Carol R. Horowitz, Lili Chan, Patricia H. Kovatch, Ira Hofer, Karandeep Singh, Lynne D. Richardson, Emmanuel Mensah, Alexander W Charney, David L. Reich, Jianying Hu, Girish N. Nadkarni

## Abstract

**Background:** Broad adoption of artificial intelligence (AI) algorithms in healthcare has led to perpetuation of bias found in datasets used for algorithm training. Methods to mitigate bias involve approaches after training leading to tradeoffs between sensitivity and specificity. There have been limited efforts to address bias at the level of the data for algorithm generation.

**Methods:** We generate a data-centric, but algorithm-agnostic approach to evaluate dataset bias by investigating how the relationships between different groups are learned at different sample sizes. We name this method AEquity and define a metric AEq. We then apply a systematic analysis of AEq values across subpopulations to identify and mitigate manifestations of racial bias.

**Findings:** We demonstrate that AEquity helps mitigate different biases in three different chest radiograph datasets, a healthcare costs dataset, and when using tabularized electronic health record data for mortality prediction. In the healthcare costs dataset, we show that AEquity is a more sensitive metric of label bias than model performance. AEquity can be utilized for label selection when standard fairness metrics fail. In the chest radiographs dataset, we show that AEquity can help optimize dataset selection to mitigate bias, as measured by nine different fairness metrics across nine of the most frequent diagnoses and four different protected categories (race, sex, insurance status, age) and the intersections of race and sex. We benchmark against approaches currently used after algorithm training including recalibration and balanced empirical risk minimization. Finally, we utilize AEquity to characterize and mitigate a previously unreported bias in mortality prediction with the widely used National Health and Nutrition Examination Survey (NHANES) dataset, showing that AEquity outperforms currently used approaches, and is effective at both small and large sample sizes.

**Interpretation:** AEquity can identify and mitigate bias in known biased datasets through different strategies and an unreported bias in a widely used dataset.

**Summary:** AEquity, a machine learning approach can identify and mitigate bias the level of datasets used to train algorithms. We demonstrate it can mitigate known cases of bias better than existing methods, and detect and mitigate bias that was previously unreported.

**EVIDENCE IN CONTEXT:** *Evidence before this study:* Methods to mitigate algorithmic bias typically involve adjustments made after training, leading to a tradeoff between sensitivity and specificity. There have been limited efforts to mitigate bias at the level of the data.

*Added value of this study:* This study introduces a machine learning based method, AEquity, which analyzes the learnability of data from subpopulations at different sample sizes, which can then be used to intervene on the larger dataset to mitigate bias. The study demonstrates the detection and mitigation of bias in two scenarios where bias had been previously reported. It also demonstrates the detection and mitigation of bias the widely used National Health and Nutrition Examination Survey (NHANES) dataset, which was previously unknown.

*Implications of all available evidence:* AEquity is a complementary approach that can be used early in the algorithm lifecycle to characterize and mitigate bias and thus prevent perpetuation of algorithmic disparities.

## INTRODUCTION

The adoption of diagnostic and prognostic algorithms in healthcare has led to perpetuation of bias against underrepresented groups. These biases exist even in algorithms trained using diverse populations, because longstanding societal biases and disparities are reflected in the data used to develop algorithms ^1^.

Attempts to mitigate bias have focused on fairness-metric based algorithm optimization after training on the data ^2–4^. Recent developments in generative algorithms have focused on amplifying representation ^5^. However, if the underlying dataset is biased, algorithms will exhibit residual unfairness or performance tradeoffs even after algorithm optimization or synthetic data generation ^6–9^. Second, many forms of implicit bias are not captured by fairness metrics or algorithm performance, and detecting these biases is difficult ^6,10^. There is increasing recognition of the need to address bias earlier in the algorithm development pipeline, when data is collected and curated or when outcomes are chosen ^11–13^.

We present AEquity, a machine learning-based method, which is used to detect and evaluate bias and perform an appropriate intervention to mitigate it using a small number of training samples. Since this technique is agnostic of the machine learning algorithm and architecture, it can be applied in a variety of settings. We first demonstrate its application to detect and mitigate bias in two scenarios wherein healthcare algorithms have been shown to be biased: 1) an algorithm used to predict healthcare costs which exhibits bias against Black patients ^10^ (healthcare costs scenario), and 2) a standard computer vision algorithm applied to chest radiographs results in selective underdiagnosis in people who are Black, Non-White Hispanic/Latino, or Medicaid ^14^ (chest radiographs scenario). Finally, we utilize AEquity to detect, characterize and mitigate a previously unreported, undetected bias in prediction of mortality with the National Health and Nutrition Examination Survey dataset (NHANES scenario)^15^. We provide an easy-to-use implementation and release this for use without any restrictions (**Supplementary Methods**).

Generally, the task can be described as: We have a subset of data containing two distinct groups distinguished by a characteristic (like race), with features x and potential outcomes y. The first task is to determine which outcome (or label) should be considered to minimize residual bias towards a group, where residual bias is after using methods after training^15^. We demonstrate optimal label selection to minimize bias using the healthcare costs scenario. The next task is to determine which samples from the dataset need to be prioritized to reduce differences in performances against, or in favor of, a group based on a characteristic. We demonstrate guided sample selection using the chest radiographs scenario and demonstrate that this works for algorithms based on both convolutional neural network and transformer architectures, which form the foundation of computer vision. Finally, we demonstrate the use of AEquity to sequentially perform label selection and guided sample selection in the NHANES scenario, where prior biases have not been reported. Thus, we demonstrate a task-, algorithm- and architecture-agnostic method for data-centric debiasing, potentially usable beyond healthcare tasks. We make it open source with documentation and a video tutorial to increase accessibility.

## METHODS

For clarity, we use “groups” to refer to various subpopulations, for example by race, gender, or insurance status. We will use “outcome” to refer to diagnostic findings, for example “pneumonia” and “edema”. A “protected group” refers to categories of people who are legally protected from discrimination under US law. It includes, among other things, age, sex, and race. Following an accepted framework ^1^, we characterize dataset biases into main types: sampling bias; complexity bias; and label bias (Figure 1). Sampling bias arises when there is a low frequency of patients with a given diagnosis from a protected group. In complexity bias, a protected group presents more heterogeneously with a diagnosis, and consequently, one group exhibits a class label with greater complexity than another group. Subsequently, even if groups are drawn with equivalent frequencies, generalization performance on the more complex group is worse than other groups for the same class. For example, this could cover a group for whom the diagnostic criteria aren’t precise due to historical reasons, a group with large translational distribution shift due to geographic differences, or a group for which there are differences in data quality. In each of these three cases, the complexity of the underlying data-distribution is expected to increase. Third, label bias occurs when outcome labels are placed incorrectly at different rates for different groups and can lead to increased misclassification errors for the affected group. For example, in published work, Black patients are more frequently labeled as requiring fewer healthcare resources despite having the same number of comorbidities because of lack of access to care ^10^.

**Figure 1.**
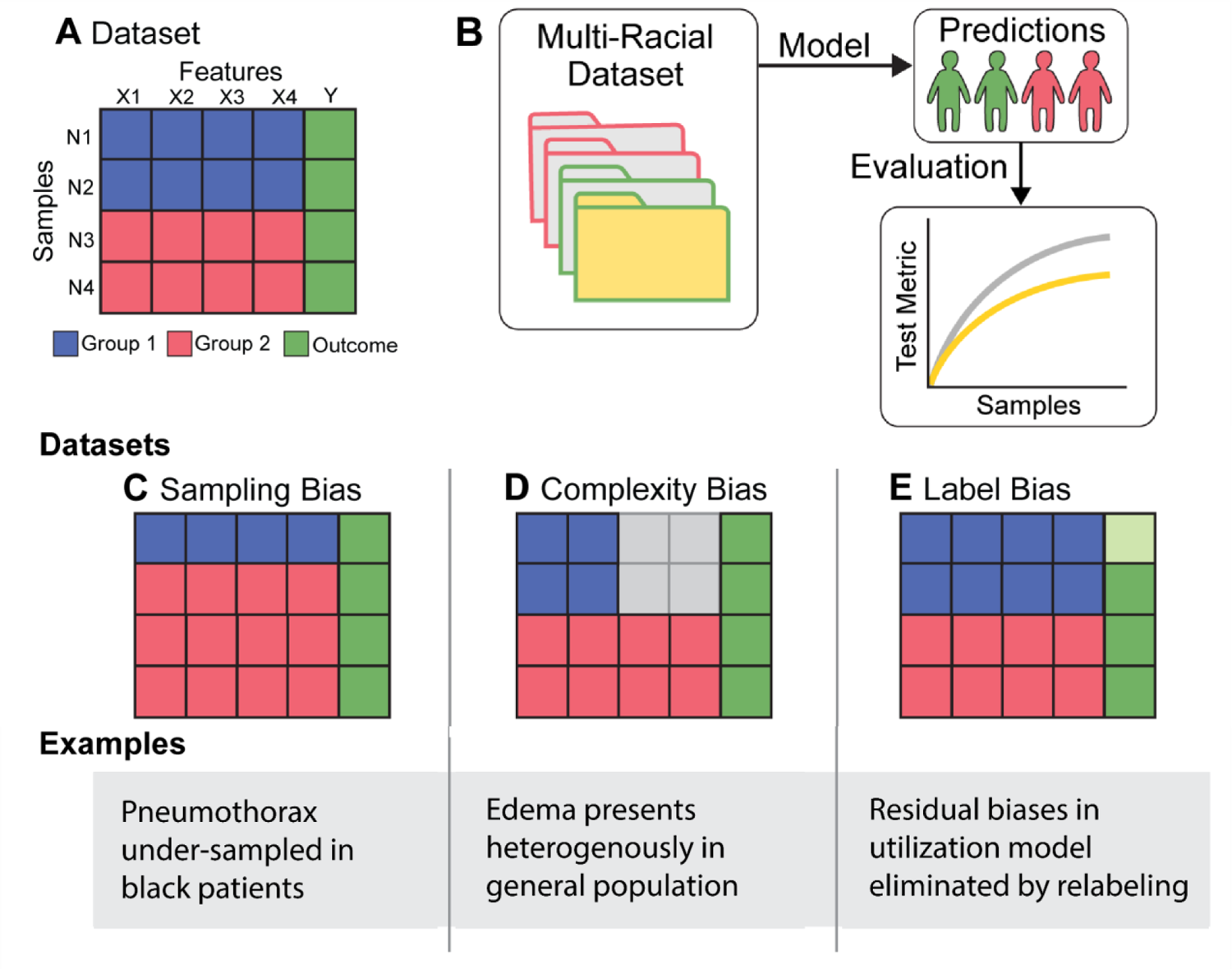
Description of Different Biases and Proposed Role of AEquity in Identifying and Mitigating Bias. A. A prototypical dataset containing samples from two groups each of which has four informative features. B. When trained in a similar manner on a more diverse dataset, the resulting model is biased because it performs worse for a particular group. C. Sampling bias occurs when one group is dramatically under-represented compared to a second group. D. Complexity bias results when one group presents more heterogeneously than a second group; for example patients with edema on chest radiographs present more heterogeneously in the general population than in each subgroup. E. Label bias occurs when mislabeling burdens one group more than another; Light green represents a higher burden of mislabeling.

We develop and validate AEquity, a machine learning-based method, that can be used for the characterization of these three biases (sampling, complexity, and label), followed by guided mitigation, in healthcare datasets. AEquity works by appending a simple compressive network to a dataset or the latent space of existing machine learning algorithms to generate a single value, AEq, characterizing an outcome with respect to a group and the priors established by the algorithm. In general, a higher AEq value for a group with respect to an outcome and a given machine learning algorithm suggests that generalization is more difficult and more or higher quality samples are needed for learning ^19,20^. Thus, AEq is a metric that is calculated at the intersection of an outcome like “pneumonia”, a group label like “Black”, and a machine learning algorithm like “ResNet-50.” AEq is reported as log_2_(sample size estimate). AEq values are only meaningful when constrained to comparing groups with the same outcome in the dataset, and interpretation of the metric is invariant to architecture of the machine learning algorithm, the compressive network or the dataset type or characteristics ^17–19^. Related mathematical work has sought to characterize the capacity of neural networks with topological data analysis and approximations of inductive bias complexity, which we do by utilizing a small number of samples to establish empirical approximations of relationships between groups ^20,21^.

To mitigate a specific type of bias, the AEq values can be used to evaluate the right composition of the dataset for training a machine learning algorithm, or for label selection without changing the dataset composition. To evaluate bias mitigation, we evaluated an extensive variety of fairness metrics (area-under-the-curve, true positive rate, false negative rate, true negative rate, positive predictive value, false positive rate, false discovery rate, and precision) and showed that AEquity can help improve these metrics over just algorithm optimization. While previous work has sought to optimize for these metrics, these works are algorithm-centric and optimization based, rather than modifications of the underlying dataset. Previous methods that have utilized information bottleneck theory make no changes to the dataset nor identify which label to best select, and therefore lead to a tradeoff between overall accuracy and fairness, which is true of any algorithm-centered approach ^22,23^. Our approach is fundamentally different – engineering the dataset used for training and selecting appropriate labels so that fairness is improved without sacrificing accuracy. While data-centric approaches in the literature have been discussed ^7,24^, state-of-the-art remains balanced empirical risk minimization ^12^, which performs at the same level as or worse than AEquity. Moreover, AEquity functions at a significantly smaller sample size (3-10% of the data). As a result, AEquity can be used early in the algorithm development pipeline and can be complementary to downstream optimizations.

### Description of AEquity based data curation

AEquity-based data curation is achieved through a multi-step pipeline (Figure 2). First, a group-balanced subset of the patient data is collected, and this data is labeled. Next, AEq values by group for each outcome are calculated. The difference in the AEq values between groups for each outcome can help us determine if label bias exists in the dataset. Fairness metrics are not sensitive measures of label bias because algorithms can learn the relationship between specific patient populations and a given outcome, even if the outcome is systemically distorted ^6,10^. However, if AEq measures are significantly different between groups for the same label, then the label may not represent the same distribution for different groups. The label with least distributional difference (smallest difference in AEq values between groups) should be selected to mitigate the bias. Next, AEq is calculated with respect to each outcome when groups are combined into a joint dataset. The relationship between individual group and joint AEq values allows us to determine if sampling or complexity bias exists, and the differences in AEq values for different patient groups for the same label are an indicator of label bias. When predictive bias is driven primarily by sampling bias in the dataset, combining groups drives the AEq value at or below the over-sampled data (**Supplementary Methods**). In sampling bias, balanced sampling from each group is sufficient to mitigate the bias because the data is equitably represented within the algorithm. When complexity bias was the only type of dataset bias, combining the groups results either in an increase in the value of AEq or an AEq closer to value that had been higher prior to the combination. In complexity bias, collecting data exclusively from the protected population, due to its relative heterogeneity compared to the over-represented group, is necessary to mitigate bias. If the algorithm exhibits residual unfairness, then the AEq will be different for two groups for the same outcome. Thus, AEquity can help with quantifying biases within outcome metrics and subsequently selecting appropriate outcomes to minimize AEq values between groups.

**Figure 2.**
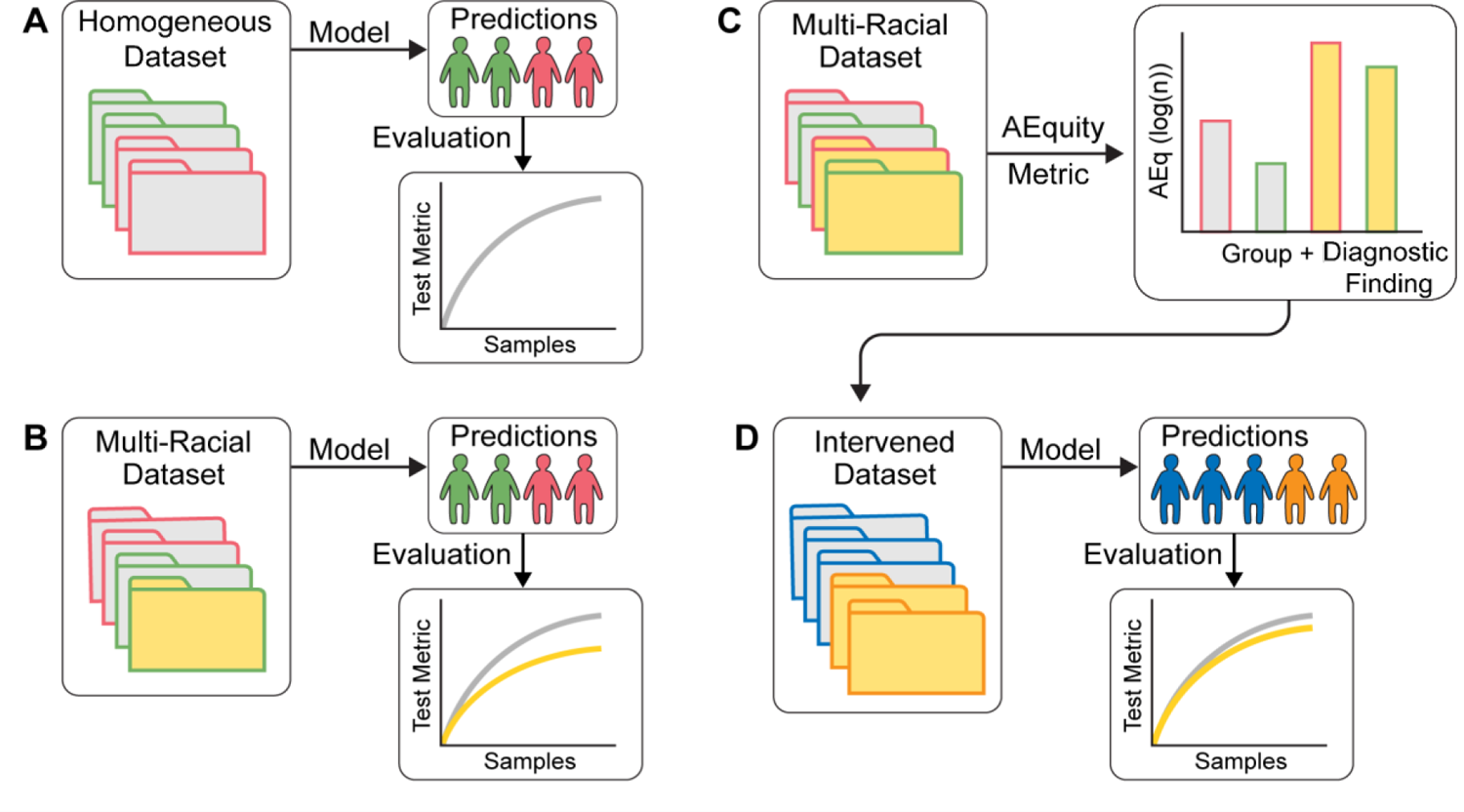
Schema Of AEquity Workflow to detect and mitigate diagnostic biases in MIMIC-CXR. A. Red – White individuals; Blue – Black individuals. B. Frequency of diagnostic findings by race. Purple – average of White and Black. C. AEquity values (log(N)) by diagnosis and race. D. Effect of data-centric mitigation intervention on bias against Black patients. E. Decrease in bias as measured by different fairness metrics in Black patients on Medicaid.

### Machine Learning and Statistical Analysis of AEquity based data curation

For all machine learning tasks, the training, validation and test set are kept independent with no overlapping patients. The training and validation sets were seeded and bootstrapped fifty times. AEquity was applied to a small subset of the training data for all three tasks (< 10%). The mean and standard errors are calculated across all fifty bootstraps and 95% confidence intervals are reported. P-values are calculated via t-tests for two-group quantitative analyses and ANOVA for multi-group quantitative variant analyses. All reported P-values are adjusted with Bonferroni correction.

When training machine learning algorithms, all algorithms are trained on an independent training set, validated on a validation set, and performances are reported on an independent test set (10%). The test set consists of held-out (non-overlapping) samples prior to any dataset modification or intervention. Training and validation sets are bootstrapped fifty times, and performance is reported on the held-out test set. Means and 95% confidence interval are reported.

## RESULTS

### Detection and Mitigation of Racial Bias from Systematic Label Distortion in Electronic Healthcare Record Datasets at Small Sample Sizes (healthcare costs scenario)

In this example, we highlight the role of AEquity when labels for a particular group are systematically distorted for a given patient population. Obermeyer et al. showed that a algorithm’s predictive power is not a sensitive metric for bias. In other words, an algorithm trained on labels that are significantly biased against Black patients has an accuracy comparable to an algorithm trained with more equitable labels. Moreover, despite equivalent predictive power, the algorithm trained on biased labels produces biased outcomes. We examined whether AEquity can detect the biased label and help select a more equitable label.

We examined a publicly available tabularized EHR dataset of 49,618 primary care patients enrolled in risk-based contracts from 2013 to 2015, from a large academic hospital, described in Obermeyer et al^1^. Data included demographics and visit information. In that paper, the authors examined an algorithm which predicted total healthcare costs for a patient as a proxy for their predicted healthcare needs. They categorized individuals at or above the 50th percentile of predicted outcome as ‘high risk’ and these patients would be screened for a care management program, whereas patients at the 95th percentile would be automatically enrolled. They found that Black patients screened for the high-risk category were significantly sicker compared to White patients in that category. This is a form of bias because it resulted in a reduced allocation of resources to Black patients in comparison with their healthcare needs.

They found that this discrepancy persisted when patients were categorized based on another predicted cost-based metric-avoidable costs. However, the discrepancy disappeared if predicted active chronic conditions were used for categorization. In short, they discuss in their paper that using a better label can mitigate bias. With AEquity, we can empirically replicate these conclusions quantitatively at small sample sizes, and subsequently guide the development of minimally biased algorithms prior to complete data collection, algorithm training and validation.

AEquity-based data curation was achieved through a multi-step pipeline. First, a race-balanced subset of the patient data was collected (n=1,024, 3% of the total available data) and this data was labeled according to each of the available labels (active chronic conditions, avoidable costs, and total costs). We calculated AEq by race (Black, White), for each risk category (high risk, low risk), for each of the three outcomes (total costs, avoidable costs, active chronic conditions) (Table 1). To determine if Black and White patients in each risk category were similar, we then calculated the difference in AEq between races, for each risk category, for each outcome.

**Table 1.** Measurement of the discrepancy between Black and White patients for each risk score in the EHR dataset using the AEquity metric. We show the AEquity metric calculated with respect to each risk metric in both the predicted low risk and high-risk population. When stratifying via avoidable costs and total costs, the AEquity metric is elevated, but when stratifying by active chronic conditions, the AEquity metric is not elevated indicating that active chronic conditions is a better choice of outcome to mitigate bias. We report 95% confidence intervals in the parenthesis.

| Risk Metric | Risk Score | AEquity Metric |
| --- | --- | --- |
| Avoidable Costs | Low Risk | 0.81 (0.71, 0.92) |
|  | High Risk | 0.41 (0.32, 0.49) |
| Total Cost | Low Risk | 0.81 (0.73, 0.90) |
|  | High Risk | 0.47 (0.36, 0.57) |
| Active Chronic Conditions | Low Risk | 0.34 (0.26, 0.43) |
|  | High Risk | 0.06 (-0.04, 0.16) |

First, the difference in AEq was statistically significant across outcome measures for the population designated as high-risk (ANOVA, P = 1.59×10^-8^). When predicted cost-based metrics (total costs, avoidable costs) were used to designate patients at highest risk, there was a significant difference in AEq between Black and White patients (Figure 3A, 3B; Table 1). For example, with total costs, the difference was 0.47 (95% CI, 0.36-0.52). Thus, Black patients predicted to be at high risk were significantly different from their White counterparts which led to inequitable resource allocation.

**Figure 3.**
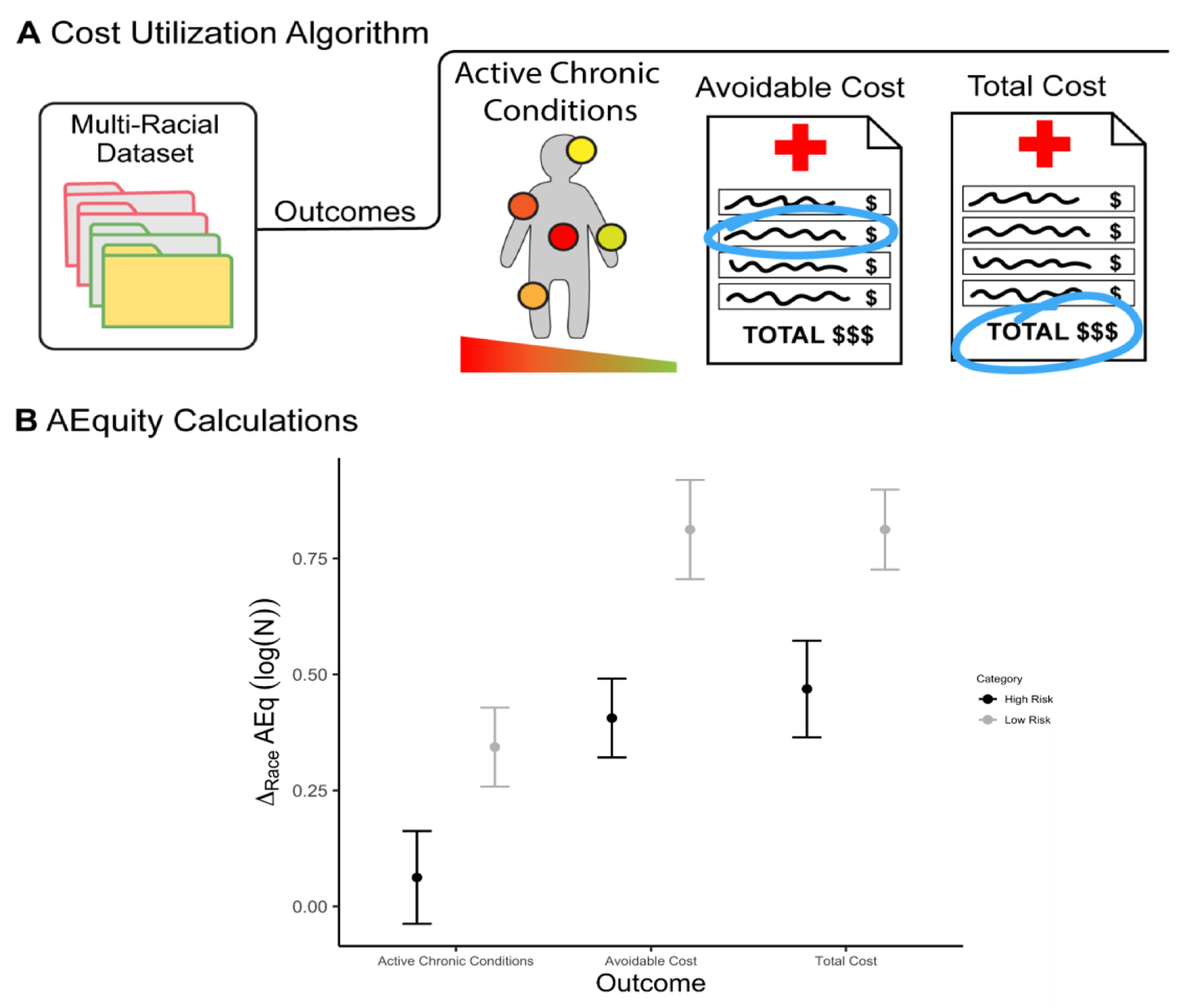
Detecting bias in prediction of healthcare needs with the AEquity metric. A. A dataset with claims data used to calculate a score based on number of active chronic conditions, and two cost-based metrics. B. Difference in AEquity values between Black and White patients for comorbidity- and cost-based outcomes, stratified by risk level.

However, the difference between Black and White patients categorized to be at high risk disappeared when using predicted active chronic conditions (instead of predicted costs) as the outcome measure; the difference in AEq was not statistically significantly different from 0 [0.06 (95% CI, −0.04 to 0.16)], indicating that Black and White patients in that risk category were similar. Thus, AEq values could *a priori* guide the choice of a better outcome measure to identify patients at high risk and thus mitigate bias.

We conducted additional analyses in patients at lower risk and found that the differences in AEq across races were all statistically significant (Figure 3B). In the low-risk group, Black patients display more heterogeneous underlying characteristics than White patients. We also observed that the differences in the AEq metric were smaller when using active chronic conditions than avoidable costs or total costs, indicating mitigation of bias for this lower-risk group despite the increased heterogeneity (Table 1). Nevertheless, the presence of differences in AEquity-score in the low-risk group may indicate that there may be some residual complexity or sampling bias even after changing the outcome metric.

### Detection at Small Sample Sizes and Mitigation of Under-diagnosis Bias in Chest X-ray (chest radiographs scenario)

We then evaluated AEquity on MIMIC-CXR, a publicly available dataset containing 377,110 chest radiographs of 65,379 patients from the Beth Israel Deaconess Medical Center between 2011 and 2016 ^25^. We also used two additional publicly available chest radiograph datasets to validate our findings – CheXpert containing 224,316 chest radiographs from 65,240 patients, and ChestX-ray14 containing 108,948 chest radiographs from 32,717 patients ^26,27^. MIMIC-CXR and CheXpert each contain 14 binary diagnostic findings, ChestX-ray14 contains 15, and all three contain an additional binary label for radiographs with no positive diagnostic finding. (Supplementary Table 1) ^26,27^. We generated a subset from MIMIC-CXR by including only postero-anterior radiographs of patients who self-identified as either Black or White, and with either one of the nine most frequently encountered diagnostic findings (atelectasis, cardiomegaly, consolidation, edema, effusion, enlarged cardiomediastinum, opacity, pneumonia, pneumothorax) or ‘no finding’. In Seyyed-Kalantari et al. ^27–29^, a convolutional neural network was trained on Chest X-rays belonging to the MIMIC-CXR dataset and evaluated on protected populations. The algorithms demonstrated significant differences in performance on protected populations, indicating bias. Subsequent work has shown that fairness methods which strive to achieve better worst-group performance (GroupDRO, JTT, CRT, DFR, etc.) do not outperform balanced empirical risk minimization (balanced ERM) ^2,12,28,29^. Thus, we compared AEquity to balanced ERM.

We followed a multi-step pipeline for AEquity-based data curation. First, we collected a balanced subset of the patient data (n=1,024, 3% of the total available data, 512 Black patients, 512 white patients) and labeled it according to each of the available labels. For each diagnostic finding, we calculated AEq by race (Black, White) and also for the joint dataset containing samples from both Black and White patients. We then used these AEq values to guide data curation - choosing either a) to group-balance with an equal number of additional samples from both Black and White patients, or b) to prioritize data collection of a single group to a total of 30,000 samples, a sample size sufficient to produce performance comparable to state-of-the-art classification algorithms (Table S2). We then demonstrated that AEquity can help analyze causes of bias at both small and large dataset sizes but becomes more precise at larger sample sizes (**Supplementary Figure S7**). We split the subset 60%-20%-20% into training samples, validation samples, and test samples, ensuring no patients overlapped, and fine-tuned two different pre-trained machine learning algorithms (ResNet-50, VIT-B-16) with PyTorch Lightning ^30–32^. First, we compared these algorithms to state-of-the-art Chest X-ray classification algorithms (Table S2). Next, we showed how AEquity can improve performance over naive data collection (Figure 4, Table 2), and benchmarked against Balanced ERM, which has previously been shown to outperform other variants of subgroup characterization such as Just-Think-Twice, GroupDRO ^30–32^ (Table 3, Figure S6). Balanced empirical risk minimization counterbalances varying population frequencies with over-weighing or over-sampling under-represented groups, and while this is a valid strategy for sampling bias, AEquity demonstrates that in many instances, groups exhibit complexity bias or label bias, which require alternative strategies. We validated against threshold-independent metrics such as area-under-the-curve as well as fairness metrics such as sensitivity, specificity, FPR, FDR, FNR in intersectional populations (Table S3). We bootstrapped each experiment 50 times, in keeping with available computational resources ^33–35^. We defined bootstrapping as sampling with replacement, but avoided sampling patients and their chest X-rays from the test dataset.

**Figure 4.**
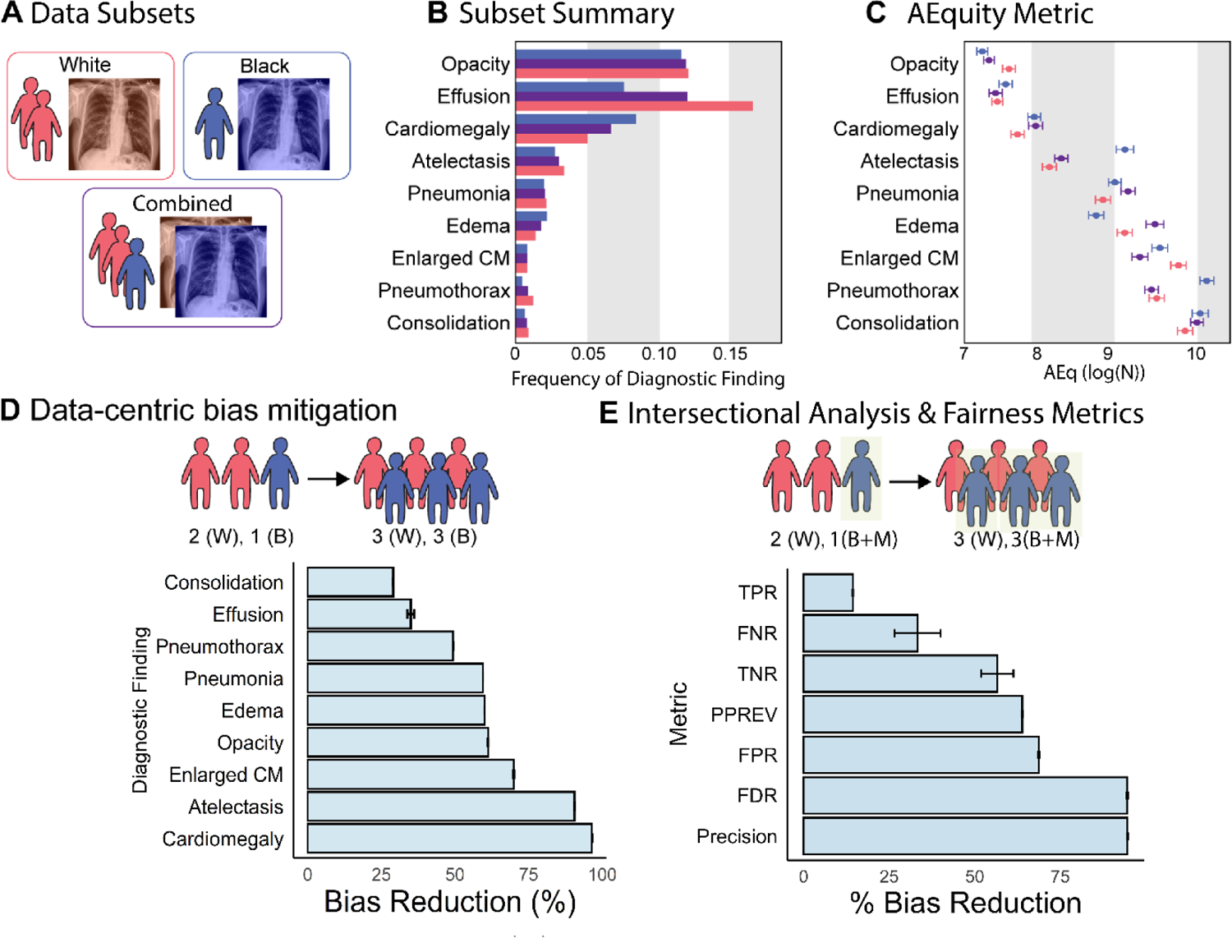
Application of AEquity to detect and mitigate diagnostic biases in MIMIC-CXR. A. Red – White individuals; Blue – Black individuals. B. Frequency of diagnostic findings by race. Purple – average of White and Black. C. AEquity values (log(N)) by diagnosis and race. D. Effect of data-centric mitigation intervention on bias against Black patients. E. Decrease in bias as measured by different fairness metrics in Black patients on Medicaid.

**Table 2.** Measurement of bias and intervention effect by diagnosis in the radiograph dataset with RadImageNet (ResNet-50 with pre-training on Medical Images) We show AUROC before and after intervention with 95% Confidence Intervals, for Black and White patients. Bias is the difference in the AUROC between Black and White patients calculated in percentages. With a dataset intervention, we reduce the discrepancy between the two groups and calculate percentage change in the discrepancy. Percentages have been rounded to one decimal place, and all other numbers have been rounded to two decimal places. * - diagnostic findings with bias against Black patients.

| Data Curation | Pre-Intervention |  |  | AEquity-Guided |  |  | Bias Reduction |
| --- | --- | --- | --- | --- | --- | --- | --- |
| Diagnosis | White Patients | Black Patients | Bias | White Patients | Black Patients | Bias |  |
| Pneumothorax* | 0.73<br>(0.72,0.74) | 0.68<br>(0.67,0.69) | 7.5 | 0.73<br>(0.72,0.74) | 0.70<br>(0.68,0.72) | 3.8 | 49.4<br>(49.3, 49.5) |
| Atelectasis* | 0.69<br>(0.68, 0.69) | 0.64<br>(0.64,0.65) | 6.4 | 0.70<br>(0.70,0.70) | 0.69<br>(0.69,0.69) | 0.6 | 90.6<br>(90.5, 90.7) |
| Edema* | 0.75<br>(0.75,0.75) | 0.72<br>(0.71,0.72) | 5.0 | 0.78<br>(0.78,0.78) | 0.79<br>(0.79,0.80) | 2.0 | 60.0<br>(59.9, 60.1) |
| Consolidation* | 0.67<br>(0.67,0.67) | 0.65<br>(0.64,0.66) | 3.8 | 0.63<br>(0.63,0.63) | 0.61<br>(0.60,0.63) | 2.8 | 29.0<br>(28.8,29.2) |
| Pneumonia* | 0.67<br>(0.66,0.67) | 0.64<br>(0.64,0.65) | 3.8 | 0.65<br>(0.65,0.65) | 0.64<br>(0.64,0.65) | 1.5 | 59.5<br>(59.4, 59.6) |
| Enlarged CM* | 0.62<br>(0.61,0.63) | 0.60<br>(0.59,0.60) | 3.6 | 0.61<br>(0.61,0.62) | 0.61<br>(0.60,0.61) | 1.1 | 69.9<br>(69.6, 70.1) |
| Cardiomegaly* | 0.76 (0.76,0.76) | 0.75<br>(0.75,0.75) | 1.9 | 0.76<br>(0.76,0.76) | 0.76<br>(0.76,0.76) | 0.1 | 96.5<br>(96.4,96.6) |
| Pleural Effusion | 0.64<br>(0.64, 0.64) | 0.64<br>(0.64,0.64) | 0.3 | 0.65<br>(0.65,0.65) | 0.65<br>(0.65,0.65) | 0.2 | 35.0<br>(33.7,36.2) |
| Opacity | 0.69<br>(0.69, 0.69) | 0.71<br>(0.70,0.71) | -2.0 | 0.69<br>(0.69,0.69) | 0.70<br>(0.69,0.70) | -0.8 | 61.2<br>(61.0,61.4) |

**Table 3.** Percentage Bias against Black patients by method in a ResNet-50 naive to medical images as measured by Area-Under-the-Curve. Balanced sampling of groups has been previously determined to be state-of-the-art across different fairness-optimizing procedures (^70^). We benchmark against empirical risk minimization procedures, showing the advantages of using AEquity over ERM.

| Label | Balanced ERM<sup>1</sup> (%) | AEquity (%) | % Improvement over sub-optimal policy |
| --- | --- | --- | --- |
| Atelectasis | 0.85 (0.76, 0.94) | **0.51 (0.42, 0.60)** | 40.07 (39.09, 41.05) |
| Pneumonia | 1.70 (1.64, 1.76) | **1.39 (1.33, 1.45)** | 24.3 (24.26, 24.34) |
| Pneumothorax | 7.11 (7.00, 7.22) | **6.42 (6.31, 6.53)** | 24.79 (24.61, 24.79) |
| Consolidation | **2.36 (2.24, 2.48)** | **2.36 (2.24, 2.48)** | 0.00 (-0.7, 0.7) |
| Edema | 2.11 (2.06, 2.16) | 2.03 (1.98, 2.08) | 3.77 (3.73, 3.81) |
| Effusion | 0.41 (0.35, 0.46) | **0.35 (0.30, 0.41)** | 12.9 (10.61, 15.21) |
| Cardiomegaly | **0.18 (-0.13, 0.49)** | **0.18 (-0.13, 0.49)** | 0.66 (0.54, 0.78) |
| Enlarged CM | 3.59 (3.42, 3.76) | **3.12 (2.95, 3.29)** | 13.1 (12.83, 13.35) |
| Opacity | 0.49 (0.38, 0.60) | **0.24 (0.13, 0.36)** | 50.3 (50.10, 50.34) |

First, we observed that a higher frequency of occurrence for a diagnosis corresponded to a lower AEq value across Black and White patients (Figure 4c). This indicates that labels with more samples, like lung opacity (AEq ≈ 7.5), generally trend towards lower AEqs, whereas labels with fewer samples such as consolidation (AEq ≈ 9.8), enlarged cardio-mediastinum (AEq ≈ 9.9) or pneumothorax (AEq ≈ 9.95) trend towards having higher AEqs. We posit that the latter group of diagnoses may have the highest potential for bias because of difficulty in generalization samples from patients not present in the training dataset. Second, we noticed that six out of the nine diagnoses, accounting for 71% of the positive samples, demonstrate higher AEq values in Black patients than in White patients. Higher AEq values indicate that, in general, algorithms built on chest X-rays from Black patients for Black patients generalize more poorly than algorithms trained on chest X-rays from White patients for the purpose of diagnosing White patients. In other words, more samples from Black patients than White patients are required to mitigate bias.

Next, we examined pneumothorax. Chest X-rays from Black individuals diagnosed with pneumothorax have significantly larger AEq values than chest X-rays belonging to White individuals with the same diagnosis (AEq_Black, Pneumothorax_ = 10.10; 95% CI: (10.01, 10.18), AEq_White, Pneumothorax_ = 9.50; 95% CI: (9.44, 9.58); P < 0.05). For the diagnosis of pneumothorax, the AEq of the joint dataset is smaller than the AEq of chest X-rays belonging to each subgroup, which implies that (AEq_Joint, Pneumothorax_ = 9.44, 95% CI: (9.35, 9.52)). Subsequently, we see that adding dataset diversity by equitably sampling from each race was associated with an improvement in classifier performance for Black individuals when compared to the naïve approach (Bias Reduction (Absolute) = 2.43 x 10^-2^; 95% CI: (2.22 x 10^-2^, 2.59 x 10^-2^); Bias Reduction (%) = 49.37%, 95% CI: (49.3, 49.5)). Second, we examined edema, where the joint AEq was higher than the AEq for either race, consistent with complexity bias. For edema, AEq_Joint_ is larger than both AEq_Black_ and AEq_White_ (AEq_Joint, Edema_ = 9.48; 95% CI: (9.37, 9.58), AEq_Black, Edema_ = 8.78; 95% CI: (8.69, 8.86); AEq_White, Edema_ = 9.12; 95% CI: (9.08, 9.20), P < 0.05).

Prioritizing data collection from Black patients better captured the group-specific presentations, and this strategy was associated with an algorithm that generalized better to Black patients when compared to a naïve approach (Bias Reduction (Absolute) = 1.95 x 10^-2^; 95% CI: (1.90 x 10^-2^, 2.01 x 10^-2^); Bias Reduction (%) = 60.02%, 95% CI: (59.98, 60.06)). Bias for each diagnostic finding in the chest radiograph dataset decreased by between 29% and 96.5% following intervention (Table 2; Figure 4D).

Finally, we demonstrated that 1) AEquity remains valid for mitigation of bias by sex, 2) AEquity-guided data curation is effective for individuals at the intersection of more than one disadvantaged group, 3) targeted data collection and curation is robust to choice of fairness metric, and 4) these results are robust when trained on transformer architectures with 86 million parameters. When evaluating sex, the joint AEq score for all 9 diagnostic findings was not significantly different from, or lower than the AEq score for females, indicating that predictive biases are primarily driven by sampling bias (Figure S1). Larrazabal et al ^12^ has previously demonstrated that group balancing is the optimal approach to mitigate gender biases in this setting. Second, we demonstrate the effectiveness of AEquity on intersectional populations, who are at greater risk of being affected by systemic biases ^36,37^, and we show that this technique is robust to the choice of fairness metrics. When we examined Black patients on Medicaid, at the intersection of race and socioeconomic status, we found that AEquity-based interventions reduced overall false negative rate by 33.3% (Bias Reduction Absolute = 1.88 x 10^-1^; 95% CI (1.4×10^-1^, 2.5×10^-1^); Bias Reduction (%) 33.3% (95% CI, 26.6-40.0)), Precision Bias by 7.50×10^-2^; 95% CI (7.48×10^-2^, 7.51×10^-2^); Bias Reduction (%) 94.6% (95% CI, 94.5-94.7%); False Discovery Rate by 94.5% (Absolute Bias Reduction = 3.50×10^-2^; 95% CI: (3.49×10^-2^, 3.50×10^-2^) (Figure S4). Finally, we demonstrated AEquity works on VIT_B_16, an 86 million parameter Vision Transformer (Figure S6). AEquity outperformed balanced ERM across every diagnostic finding when measuring bias and resulted in reductions of up to 55% in bias compared to the originaldataset (Table S5).

Notably, in four of the nine diagnostic findings (opacity, effusion, edema and enlarged cardiomediastinum), analysis of AEquity values resulted in the curation of a dataset reflecting the underlying empirical group frequencies. However, in the remaining five (cardiomegaly, pneumonia, pneumothorax, and consolidation), AEquity led to a dataset that prioritized one group, outperforming balanced ERM for the mitigation of bias^38^.

We do not examine label bias due to unavailability of gold-standard labels besides those already present in publicly available dataset. However, Zhang et al have previously shown that, at least for the ‘No Finding’ label in the chest radiograph dataset, the prevalence of incorrect labeling did not vary significantly by race and sex, but that it was more prevalent in the subgroup of patients over age 80 ^38–40^.

### Detection of Previously Unknown Bias in the National Health and Nutrition Examination Survey for predicting all-cause mortality (NHANES scenario)

The National Health and Nutrition Examination Survey (NHANES) data was collected between 1999 and 2014, and includes demographic, laboratory, examination and questionnaire features. The NHANES dataset has been used for a wide range of applications including the measurement of the population prevalence of metabolic syndrome, demographic trends in major depressive disorder, and associations between factors such as inflammation and autoimmune diseases ^39–41^. Machine learning has been used on the NHANES dataset to predict coronary heart disease, hypertension, and all-cause mortality ^42–44^.

The prediction of all-cause mortality is an important task because all-cause mortality is a commonly chosen outcome in epidemiological studies ^45^. All-cause mortality can be used as a metric for health disparities, to guide public health interventions, and to affect health resource allocation ^46–48^. Therefore, detecting, characterizing, and mitigating biases in the prediction of all-cause mortality can potentially affect large scale epidemiological interventions. To evaluate biases in the prediction of all-mortality in the NHANES dataset, we pre-processed the data (N=47,261) and selected variables as described in past work by Qiu et al ^44^. To detect potential biases, we calculate AEq values at small sample sizes (N=400) for Black and White individuals, and all-cause mortality for different follow up periods in the NHANES dataset (Figure 5A). Next, we qualitatively and quantitatively analyzed the AEquity values to identify the types of biases in the dataset. Finally, we used AEquity to guide data-centric intervention, and compared this intervention with simple algorithmic calibration and balanced ERM. All experiments were bootstrapped 50 times with the Pytorch library.

**Figure 5.**
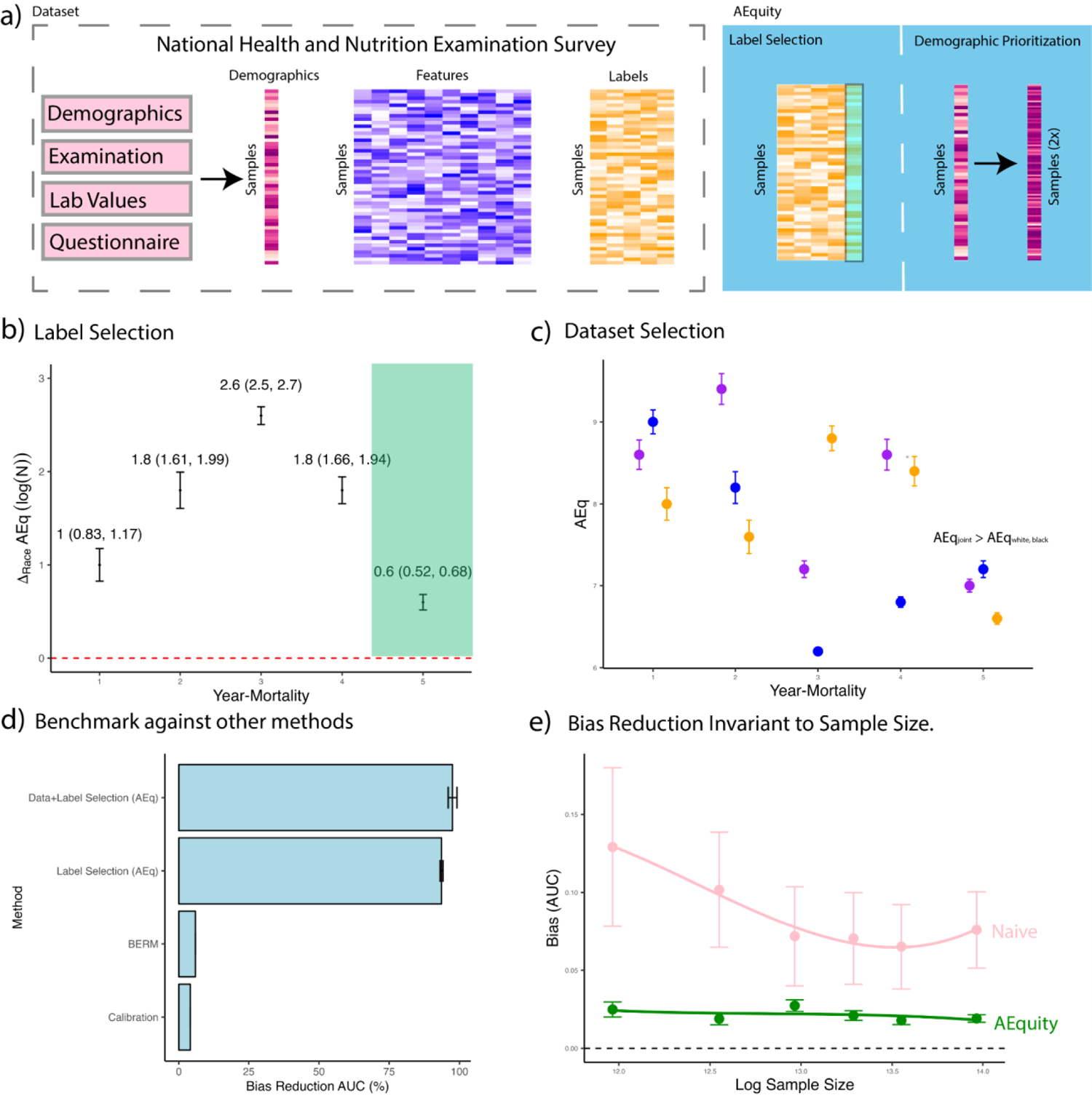
Application of AEquity to Detect, Characterize, and Mitigate a Previously Undetected Racial Bias in Mortality Prediction on a Commonly Used Dataset. A. Schematic representations of AEquity applied to label selection and demographic prioritization of bias in the National Health and Nutrition Examination Survey (NHANES) B. AEquity reveals that the least biased label for mortality prediction occurs at 5-years because the groups have similar levels of complexity. C. AEquity determines that 5-year mortality has complexity bias because the joint AEq exceeds the AEq value of both black and white patients. D. AEquity outperforms other modalities of bias reduction, namely Balanced Empirical Risk Minimization (BERM), and Calibration. E. AEquity demonstrates that bias reduction is invariant to sample size.

First, we evaluated for label bias by calculating the difference in AEquity score between Black and White patients for all-cause mortality at different follow-up periods (Figure 5B). AEquity scores for Black and White patients are least significantly different at 5-year mortality (AEq_Difference, 5-year-mortality_ = 0.60; 95% CI: (0.52, 0.68); AEq_Difference, 1-year-mortality_ = 1.00; 95% CI: (0.83, 1.17); *P < 0.05*). Next, we evaluated for complexity and sampling bias based on 5-year-mortality and race (Figure 5C). The AEq value for the joint distribution was significantly larger than the AEq values for either Black or White patients, signaling complexity bias (AEq_Joint, 5-year-mortality_ = 7.00; 95% CI: (6.93, 7.07); AEq_White, 5-year-mortality_ = 7.00; 95% CI: (6.93, 7.07); AEq_Black, 5-year-mortality_ = 6.6; 95% CI: (6.53, 6.66); *P < 0.05*).

Based on the results from AEquity, we curated a dataset with prioritized data from Black patients, trained a gradient-boosted tree to predict 5-year-mortality, and compared its fairness metrics to that of a gradient boosted tree trained to predict 1-year-mortality with a dataset sampled based on population prevalence. First, AEquity significantly reduced bias as measured by the difference in area-under-the-curve (AUC) between Black and White patients, and outperformed both calibration and balanced ERM (Figure 5D). Second, label selection and dataset prioritization had synergistic bias mitigation effects (Figure 5D). Third, AEquity’s bias mitigation relative to training with random sampling from the original dataset is invariant to sample size (Figure 5E).

## DISCUSSION

Algorithmic bias is one of the major challenges to the adoption of AI in healthcare and is a focus for regulation. For example, the Good Machine Learning Practice guidance from the FDA and EMA emphasizes the importance of ensuring that datasets are representative of the intended patient population ^49^. However, this is not sufficient because significant bias can result from the choice of outcome or differences in group complexity even when datasets are diverse. AEquity, the machine-learning based technique described in this paper, can help characterize and mitigate bias early in the algorithm development lifecycle.

The responsibility for ensuring algorithmic equity is being placed on algorithm developers and institutions. The US Department of Health and Human Services has proposed a rule under Section 1557 of the Affordable Care Act to ensure that “a covered entity must not discriminate against any individual on the basis of race, color, national origin, sex, age, or disability through the use of clinical algorithms in decision-making”. More recently, an executive order has placed at least some of the liability for algorithmic bias on developers and deployers of these algorithms ^50^. Therefore, the need to develop tools to identify and remediate potential bias in healthcare datasets is urgent from both ethical and regulatory perspectives.

The algorithm development pipeline involves task identification, data collection, data curation, training, validation, and l deployment, each of which plays an important role in whether it adds value to the healthcare system or propagates disparities that it seeks to address ^51–54^. The ability of this approach to quantify and address bias at small sample sizes makes it an attractive addition to the toolset, akin to sample size determination but for bias ^54^. Nevertheless, data-centric curation is only one of many approaches in which bias can be mitigated ^55^.

Currently, the most prevalent solution for bias mitigation with data-centric approaches is the comparison of predictive power across protected groups and balanced ERM. We have demonstrated the application of AEquity to datasets where biases have been confirmed and described in high-quality peer reviewed studies, but application of the current state-of-the-art method falls short. Algorithms predicting all three labels in the healthcare costs scenario of Obermeyer et al have similar predictive power, but two of the three are biased, demonstrating that equal predictive power is not a reliable indicator of the absence of bias. AEquity can identify the biased labels by examining a small subset of the data. Second, we demonstrated in the chest radiograph scenario that the optimal mitigation of bias sometimes requires prioritization of data from the minority population and at other times, it requires a larger, more balanced dataset.

AEquity can help distinguish these two scenarios early in the data collection process, because it only requires a small amount of data for analysis. Third, we use AEquity to describe and mitigate a previously undetected and unreported bias in all-cause mortality prediction based on data from the National Health and Nutrition Examination Survey (NHANES). Applying AEquity to the NHANES dataset can detect the bias at small sample sizes and help optimize label selection and dataset curation to mitigate bias across a range of sample sizes.

Data-guided strategies face challenges due to the lack of representation in many datasets ^56,57^. Moreover, once a gap has been identified, targeted data collection of protected populations raises ethical concerns given the long history of mistrust ^58–63^.

Nevertheless, acknowledging the existence of these biases through quantitative measures can build trust, a crucial step towards mitigating some of these biases at a grassroots level ^64,65^. Investigating the reason why AEq scores are different between different populations may provide key insights into underlying mechanisms of racial inequities ^66,67^. More formally, US regulatory guidelines require that AI algorithms in healthcare not be biased against protected classes ^68^. The collection of appropriate data to test this against protected classes is obligatory even if it is difficult. AEq can help guide sampling strategies early in the data collection process because it needs relatively few samples.

Our examination of AEquity builds on previous work examining bias in healthcare. As with many machine learning approaches and clinical studies, AEquity is empirical, and no theoretical proof exists. In past work, we highlighted that the sample size at which the training loss curve of an autoencoder converges corresponds to the sample size at which a training loss curve for a neural network classifier trained on the same data converges. We showed that this holds in a variety of computer vision datasets and on latent spaces belonging to a variety of structures, which included varying the number of classes, number of informative features, number of total features, number of class-specific subgroups, and the relationships between different variables within the dataset ^68,69^. In this work, we build on that approach to the problem of bias mitigation in healthcare. Here, we do not use it to predict sample size for training a neural network. Rather, we show that it can be used as a simple marker of the learnability of data subsets and to further guide dataset intervention which can reduce bias of classifiers trained on the data. However, because no theoretical proof exists, its performance should be verified when applying it to a new setting. Nevertheless, the demonstration of AEquity in multiple state-of-the-art algorithms, multiple tasks and multiple metrics provides strong validation for its use in practice.

While our results are promising, there are some limitations to AEquity and directions for future inquiry. First, we primarily focus on three case examples – underdiagnosis, under-allocation of resources and unreported bias for outcome prediction. However, validation of AEquity on a broader scale across different phenotypes, protected groups, data modalities and algorithms types are essential. Second, AEq in its current form focuses primarily on classification tasks. A growing number of algorithms, however, are: a) using various forms of regularization and attention to produce more informative latent spaces ^69,70^; b) being used for generation rather than simply classification or regression ^71^ and c) are multi-modal ^72^. Extending AEq to these latent spaces and generative algorithms may help investigate and mitigate bias in those settings. However, generative algorithms are not substitutes for appropriate data curation, ^5^ and generative algorithms themselves can contain biases ^73–75^. For example, the use of diffusion algorithms in conjunction with data-centric strategies may yield compounding effects ^5^. Third, the labels for the MIMIC-CXR dataset were obtained using the CheXpert system of natural language processing of radiology reports, which is known to have a substantial and varying amount of mislabeling for different diagnoses ^69^. Results from Zhang et al suggest there is no statistically detectable label bias by race in MIMIC-CXR for the ‘no finding’ label ^72^. Fourth, we cannot exclude the possibility of label bias related to the rate of mislabeling being different for different groups for other diagnoses. AEquity can help choose a less biased label, but the cost of labeling healthcare data often precludes the availability of alternative, high-quality labels. The application of frameworks for dataset development which include expert labeling of the most informative samples may help overcome this problem ^76^. We also posit that identification of the type of bias can assist in identifying the other origins of bias. For example, AEquity can help characterize the bias as primarily complexity-related, which may lead an investigator to hypothesize that it may be due to more advanced disease due to lack of access to care. However, we concede that AEquity cannot confirm that lack of access to care is indeed the cause. Similarly, in the case of label bias, one may hypothesize that structural racism may contribute to the decision to focus on labels which perpetuate inequities. However, a method like AEquity, which looks at patient-level predictors and outcomes, is not expected to be able to confirm such a hypothesis.

In summary, we present AEquity, a task-, algorithm-, and architecture-agnostic machine learning based metric that may be valuable for disentangling and quantifying various types of biases at the dataset level. We demonstrate the application of AEquity to guide specific, effective mitigation measures in datasets where biases have been confirmed and described in high-quality peer-reviewed studies. We demonstrate its robustness by applying it to different datasets and algorithms, intersectional analyses and measuring its effectiveness with respect to a range of traditional fairness metrics. Finally, we use it to detect and mitigate label and complexity bias in a dataset where bias had not been described before. Although we demonstrate its utility in the urgent use case of algorithmic bias in healthcare, this approach can be used to address biases across the spectrum of algorithmic applications.

## Data Availability

The publicly available images can be access via the PyTorch library (torchvision —Torchvision main documentation (pytorch.org)). The MIMIC-CXR version 2.0 dataset is publicly available by registration at https://physionet.org/content/mimic-cxr/2.0.0/. The CheXpert dataset is publicly available by registration at https://stanfordmlgroup.github.io/competitions/chexpert/. The NIH-CXR dataset is publicly available at https://nihcc.app.box.com/v/ChestXray-NIHCC. The dataset used in the Obermeyer analysis is available in the supplement (Dissecting racial bias in an algorithm used to manage the health of populations | Science). The code for the analyses are available here (Nadkarni-Lab/AEquitas: Deep Learning Based Metric to Mitigate Dataset Bias (github.com), and the instructions for usage are included in the supplement.

## Acknowledgements

The authors would like to acknowledge Jill Gregory for support in illustrating the figures and the staff at the Minerva High Performance Computing Cluster for maintaining the servers used to run the experiments.

## Author contributions

Conceptualization: FFG, ASS, GNN Methodology: FFG, ASS, GNN Investigation: FFG, ASS, GNN Visualization: FFG, Funding acquisition: GNN, Project administration: ASS, GNN Supervision: DLR, GNN, AWC, Writing – original draft: FFG, ASS, GNN, Writing – review & editing: LL, CRH, LC, PHK, IH, KS, AWC, JH

## Supplement

### Materials and Methods

#### Background on the AEquity metric

AEquity is rooted in information bottleneck theory. The core idea of information bottleneck theory is that a deep learning classifier mapping input X to output y does so by retaining the relevant information and eliminating the irrelevant information through a hidden layer h^1^. In contrast, an autoencoder reconstructs X, i.e., maps input X to input X through a hidden layer h and must retain all meaningful information. If y semantically encodes all the meaningful aspects of X, then the information loss between X and y through h in the classifier is minimal. The information, therefore I(X;y) is approximately equal to I(X;X) at all sample sizes, and rate of learning for the autoencoder and the generalizability of a deep learning model is similar^2^. Information flow is invariant to autoencoder architecture and loss, which makes this technique generalizable across different hyper-parameters^3^.

This approximation can be utilized for sample size determination for deep learning models. A standard practice in traditional experimental design, sample size estimation takes place prior to data generation, and describes the minimum number of samples required to observe an effect of an independent variable on an outcome. The deep learning equivalent is the minimum convergence sample, defined by ^73^, which describes the minimum number of samples needed for an algorithm trained on that dataset to start generalizing to unseen data. If f_n_ is a deep learning algorithm that maps an input X to h to y trained on n samples where h is the latent representation, then the minimum convergence sample *n* is the number of training samples such that E[AUC (fn(X_test_,y_test_)] > 0.5. In practice, this number cannot be directly observed without prospective validation because the test set, by definition, is not available until training is complete. It can, however, be approximated using an autoencoder, which we define as g_n_: X to h to X, where h is the latent representation. The minimum number of samples *n* such that E[g_n_(X_train_, X_train_)] < 1.0 is defined as the minimum convergence sample estimate. Above the minimum convergence sample, Gulamali et al have shown that the decrease in autoencoder loss is proportional to the gain in generalizability as measured by area-under-the-curve metric^4^. The minimum convergence sample estimate remains a valid approximation of generalizability across model architecture, hyperparameter selections, dataset dimensions and dataset complexity.

#### Bias is marked by differences in generalization performance by group or class

Consequently, we examined the MCSE metric on subsets of the data consisting of individual groups and classes. The resulting *AEquity* metric is defined as

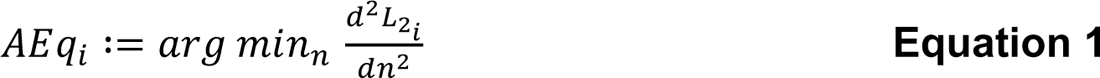

where *L*_2i_ is the *i*^th^ class/group-specific loss curve for a network trained on samples drawn from the entire dataset. A smaller *n* tends to indicate a better generalization performance, whereas a larger *n* indicates poor generalization performance when compared to other classes.

#### Code Availability

The code for the analyses is available at (removed for double-blind peer review) The steps to use the repository are listed below.

1. Install requirements. These requirements are publicly sourced such as sklearn torch scipy torchvision pandas pip install requirements.txt
2. Setup configuration in a config.yaml file. data_path: ./data/custom_data.tsv # Contains path to data. Contains independent variables, demographics, and outcome variables. demographics_col: demographics # Name of demographics variable in data_path. outcome_cols: outcome_1 # Name of outcome variable in data_path exclude_cols: None # name of columns to exclude if there are extraneous columns. out_data: ./output/data.tsv # Output directory for AEq analyses. bootstraps: 10 # Number of bootstraps. 30-50 is typically recommended for resolution at 5000 samples. start_seed: 42 # Seed experiments. input_dim: 149 # Number of independent columns in data_path max_sample_size: 5000 # Max sample size to calculate from. Usually only require 128-512 samples. root_dir: ./weights # Root directory to output weights, and other output files.
3. Run the measure and mitigate experiments. python measure_disparity.py --config config.yaml python mitigate_disparity.py --config config.yaml Additional configurations are built for more complex models (AlexNet, ResNet, EfficientNet), but require a custom dataloader. Custom dataloders can be built by modifying the cnnMCSE repository. See the cnnMCSE repository for more details.

## Materials

### Algorithm Implementation for Medical Imaging

We have utilized three large public chest radiograph datasets in this study: MIMIC-CXR, CheXpert, and ChestX-ray14 (**Table 1**). The MIMIC-CXR dataset was collected from Beth Israel Deaconess Medical Center (Boston, MA, United States) between 2011 and 2016, the CheXpert dataset was collected from Stanford Hospital (Stanford, CA, United States) between October 2002 and July 2017, and the NIH ChestX-ray14 dataset was collected from the NIH Clinical Center (Bethesda, MD, United States) between 1992 and 2015. We focus on diagnostic labels with sufficiently high frequencies.

A standard practice in deep learning on medical imaging classification tasks is the use of transfer learning – utilization of a pre-trained model such as AlexNet which is then fine-tuned on another dataset for a given task. We employ transfer learning baseline architectures and fine-tune the classification block to generate our classifiers, and the encoder-decoder blocks to generate our autoencoders. We seed each experiment with PyTorch, bootstrap experiments at 50x and report the Area Under the Curve Metric on a held-out test dataset (AUC) for our classifiers and the L_2_-norm for the autoencoders. Code is provided as an attachment.

To compute the AEq values, we utilized standard machine learning practices to generate an autoencoder with tunable weights and biases. We generated joint datasets by merging a balanced sample of chest X-rays belonging to each race, controlling for sampling bias across different groups. We adapted a pre-trained ResNet-50^77^, which has 23 million parameters for the convolutional and max pooling layers. We replaced the linear layers with an autoencoder with a latent space h = 2, selected via a grid-search based method. The autoencoder loss was trained via mean-squared error or L_2_-norm. The network was trained via Adam optimization, with a learning rate determined by a simple grid search. All experiments were bootstrapped fifty times, and error bars represent 95% Confidence Intervals. The data-loaders and networks classes were generated with PyTorch. All experiments were trained on a single NVIDIA A100 GPU using the CUDA toolkit backend. All values and 95% confidence intervals are reported. P values are unpaired t-tests for two groups, and ANOVA for multiple groups.

We used a pretrained ResNet-50 model to generate encodings for the Chest X-ray. We added three dense layers and a SoftMax layer to the top of the model to generate a single-class classifier. We used a simple grid search with values ranging from 10^-5^ to 10^-1^ for the learning rate and 4 to 256 for the batch size. After the hyperparameter optimization, we fine-tuned the dense layers and classification layers to generate classifications on a training set that was guided by AEquity values in comparison to naïve dataset collection. For example, in the case of the pneumothorax, the AEquity value dictated a diversification of the dataset, which was a 50-50 split between Blackand white patients, whereas in the case of edema, the AEquity value required population prioritization, which meant collecting samples belonging to Black patients exclusively.

The train-validation-test split was 60-20-20, with the size of the training dataset being 30,000 samples from the MIMIC Chest-X-ray dataset. We trained for a maximum of 30 epochs and employed early stopping on validation area-under-the-curve metrics. For each experiment, we bootstrap the train and validation datasets with different seeds, and show that our results for generalizability are robust to sampling. The train, validation and test set, which consist of mutually exclusive samples, are bootstrapped fifty times and 95% confidence intervals are reported.

We take a data-centric approach with simple training of a ResNet-50 pretrained model and achieve similar performance to state-of-the-art approaches which use self-supervision and knowledge distillation, label dependencies, hand-crafted features, location aware dense networks. We report the results from the knowledge distillation paper below, which was benchmarked against the latter methods (**Supplementary Table S2**).

We define bias as the discrepancy between generalization performance (as measured by the area under the receiver operating characteristic curve) for two groups, for example, White and Black patients. We choose this over traditional fairness metrics like precision or false negative rates because AEquity works directly with the dataset and doesn’t concern itself with model calibration. We later show empirically that bias mitigation with AEquity is robust to the choice of fairness metric (i.e., bias as measured by traditional fairness metrics is mitigated as well).

We calculate % reduction in bias with the following steps. First, we calculate the bias without a data-centric intervention.

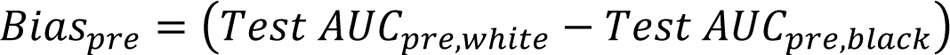

Second, we calculate the bias with a data-centric intervention.

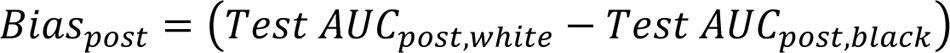

Third, we calculate the reduction in bias as a percentage of the original bias.

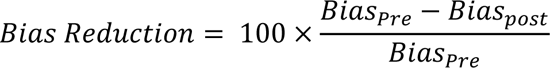

We report the reduction in bias for both diversification and population prioritization.

Algorithm Implementation for Cost-Utilization Analysis

In the data provided by (*6*), the data frame has two elements.

1. Y_it_: Outcome – Avoidable Cost, Total Cost or Comorbidity Score.
2. X_i,t-1_: The data collected by the patients insurer over the year *t-1*.

a. Demographics (age, sex excluding race)
b. Insurance type
c. ICD-9 diagnosis and procedure codes
d. Prescribed medications.
e. Encounters
f. Billed Amounts.

From these, 149 features which include biomarkers, demographics and individual datasets are generated. The output feature is either the total cost, avoidable costs due to emergency visits and hospitalizations, and health which measures how many chronic conditions are flaring up and driving utilization. Those whose metrics exceed a lower threshold, the 50th percentile, are typically referred to their primary care physicians, who are provided with additional metrics about the patients and prompted to consider whether they would benefit from enrollment. We assign them to the high-risk category.

To compute the AEquity values, we utilized standard machine learning practices to generate an autoencoder with tunable weights and biases. We generated combined datasets by merging a balanced sample of electronic health record-derived data belonging to each race, controlling for sampling bias across different groups. The tabular nature of the dataset requires a fully connected network encoder and decoder, which was trained via Adam optimization, with a learning rate determined by a simple grid search. All experiments were bootstrapped fifty times with different seeds that initialized different weights with Xavier initialization. The error bars represent 95% confidence intervals. All experiments were trained on a single NVIDIA A100 GPU using the CUDA toolkit backend. All values and 95% confidence intervals are reported. P values are unpaired t-tests for two groups, and ANOVA for multiple groups.

We provide a notebook demonstration and YouTube tutorial on how to use this method for any tabularized dataset, using the EHR dataset as the example (removed for double-blind peer review). Selecting an informative label on the electronic health record dataset can improve test AUC by 0.11 (ΔAUC = 0.109; 95% CI: (0.106 – 0.112); *P* < 0.05) on Black Patients by decreasing false negative rate and increasing precision.

## Supplementary Results

### Replication on Sex

In this section, we demonstrate the potential of using the AEquity metric to understand biases based on sex (Figure S1). For the MIMIC-CXR dataset, we notice that while most diagnoses (5/9) have a higher AEquity value for females over males, the labels that appear at higher frequencies such as opacity and effusion tend to have a higher AEquity value for males rather than females. However the joint AEquity values for these labels – opacity and effusion – tend to be lower than the AEq on individual groups, which may mitigate the biases of these labels in practice. In the CheXPert dataset, we see that only three labels appear to have statistically significant differences in AEquity values between males and females – cardiomegaly, enlarged cardio-mediastinum, and consolidation. Two of these labels have higher AEquity values for females (cardiomegaly and enlarged cardio-mediastinum), and one has a higher AEquity value for males (consolidation). In the NIH dataset, we see that while females have a lower AEquity value across the in five out of the seven diagnoses, the joint AEquity values are significantly larger in four of these, which means that training on a joint dataset may perpetuate some of the biases, and it makes more sense to further prioritize collection of data on female individuals.

### Replication on Age

Next, we look at the effect of age on AEquity values (Figure S2). In the MIMIC-CXR, we see that an older age corresponds to a higher AEquity value in three of the diagnoses – enlarged cardiomediastinum, pneumonia and pneumothorax, and a younger age corresponds to a higher AEquity value for edema and cardiomegaly. In the ChestX-ray14, we see that generally AEquity is generally balanced with a higher AEquity value for younger individuals in two out of the seven labels (opacity and pneumothorax). In the CheXPert data, we see that age is relatively balanced with a higher AEquity value in two out of the nine diagnoses for the younger age – pneumonia and effusion.

### Investigations and Interventions based on Medicare Status

We sought to validate our interventions in comparisons of insurance status (Figure S3). First, however, we investigated the coherence between age-related biases, as measured by Medicare status, and the race-related biases that we previously measured. We wanted to see if the impacts of age and race on biases, measured by AEquity, are coherent or dis-coherent.

We draw data from individuals with Medicare and those individuals with private insurance in Whites only, to control for the effects of race on data distribution observed above. We see some differences in distribution between the privately and Medicare-insured individuals. For example, chest radiographs labeled with pneumothorax in individuals with Medicare have a significantly higher AEquity value than those in individuals that are privately insured (AEq_Medicare, Pneumothorax_ = 9.89; 95% CI: (9.80, 9.90), AEquity_Private, Pneumothorax_ = 8.71; 95% CI: (8.64, 8.78); *P < 0.01*) (**Fig S3(a), 3(b)**). Moreover, the joint AEquity value is strictly in between the two different insurance status, which may suggest that there is some but not all overlap between the two distributions (AEquity_Joint, Pneumothorax_ = 8.95; 95% CI: (8.87, 9.04)). This difference was observed in racial bias experiments, which may suggest that the differences in distribution are somewhat captured by inequities in care that are overlapping between Black individuals and White individuals that are insured on Medicare. However, the joint AEquity values of the datasets merged by racewere slightly lower than either of the individual group’s AEquity values, which indicates that Blacks and Whites had overlapping distributions (AEquity_Joint, Pneumothorax_ = 9.44; 95% CI: (9.35, 9.52), AEquity_Black, Pneumothorax_ = 10.1; *95% CI*: (10.01, 10.18), AEquity_White, Pneumothorax_ = 9.50; *95% CI*: (9.41, 9.57), *P > 0.05*). When stratifying by insurance status, the joint AEquity is between the two individual insurance status, which indicates that there is a significant complexity bias in the distribution of pneumothorax when stratifying by privately and Medicare-insured individuals.

We see the opposite trend in chest radiographs labeled with a diagnosis of atelectasis where privately insured individuals have a significantly larger AEquity value than individuals on Medicare (AEquity_Private, Atelectasis_ = 8.82; *95% CI:* (8.72, 8.90), AEquity_Medicare, Atelectasis_ = 8.08; 95% CI: (8.00, 8.15); *P < 0.05*). The joint AEquity value is between the two distributions, which indicates a non-overlapping data distribution (AEquity_Joint, Atelectasis_ = 8.59; 95% CI: (8.50, 8.67)). In contrast, Black individuals have an AEquity value higher than White individuals for this specific diagnosis, which indicates that this trend cannot be solely explained by systemic disparities in healthcare (AEquity_Black,Atelectasis_ = 9.12; 95% CI: (9.01, 9.22), AEquity_White, Atelectasis_ = 8.22; 95% CI: (8.13, 8.30), *P < 0.05*).

Next, we highlighted that our interventions, which initially were conducted on race, are valid to mitigate bias as measured by Insurance status (**Fig S3(c), S3(d)**). or effusion, AEquity_joint_ is smaller than AEquity_private,_ AEquity_Medicare_. (AEquity_Private, Effusion_ = 8.08; 95% CI: (8.00, 8.16), AEquity_Medicare, Effusion_ = 7.81; 95% CI: (7.73, 7.89), AEquity_Joint, Effusion_ = 7.65; 95% CI: (7.56, 7.73)). In the case where the AEquity value requires us to increase dataset diversity, we add samples to the training dataset from each group equally – drawing equally from patients with and without private insurance. In the case where the AEquity value recommends population prioritization, we add samples exclusively from patients not on private insurance. We compare both to the naïve approach of data collection. For effusion at the largest evaluated sample size, there was a 0.02 increase in test AUC on chest X-rays belonging to individuals with Medicare following the intervention, (AUC_pre_ = 0.79; *95% CI*: (0.79, 0.80) vs. AUC_post_ = 0.83; *95% CI*: (0.82, 0.84) n = 8,192). For pneumothorax, AEq_joint_ is larger than AEquity_private_ but smaller than AEquity_Medicare_ (AEquity_Joint, Pneumothorax_ = 8.96; 95% CI: (8.87, 9.04), AEquity_Private, Pneumothorax_ = 8.71; 95% CI: (8.64, 8.78), AEquity_Medicare, Pneumothorax_ = 9.89; 95% CI: (9.80, 9.99), P < 0.05). Population prioritization leads to a 0.06 increase in test AUC for pneumothorax on Chest X-rays belonging to individuals with Medicare (AUC_Pre_ = 0.59; 95% CI: (0.59, 0.60) vs AUC_Post_ = 0.65; 95% CI: (0.65, 0.66), P < 0.05 n = 8,192)

### Intersectional Analysis on Black Patients and Age, Socioeconomic Status, and Sex

In this analysis, we investigate the intersection of race and age, socioeconomic status and sex (**Fig S4**). We run AEquity analysis on datasets that intersect between different under-represented groups including age, socioeconomic status and sex. First, we notice that for age, we compared patients that were between the ages of 20-40 and the ages of 40-60. For the analysis on the intersection of age and race, we see that AEq value is significantly higher for the older group in 5 out the 9 positive labels, versus higher for the younger group in 2 out of the 9 positive labels. The “No finding” label is higher for patients in the younger demographic, which may be explained by significant more variability in patient presentation where X-ray may have been ordered as a rule-out diagnosis rather than to specifically attribute a diagnostic cause. However, we notice that the joint is lower than that for older individuals for the labels Opacity, None, Pneumothorax, Edema and Atelectasis, which may mean that training on a joint dataset may mask some of potential biases we could have seen between the groups.

Second, we investigate the role of the intersection between sex and race. We analyze the chest X-rays in Blackmale patients and Blackfemale patients. We previously saw that white female patients generally have a higher AEquity value than their male counterparts in the MIMIC Chest X-ray dataset This trend also holds true in Black patients. We see that in four of the 9 labels, AEquity values are significantly higher for females than males, whereas we see that Blackmale patients have a significantly higher AEquity in three of the 9 labels. However, in these three labels, we see that the joint AEquity is larger than AEquity values of the individual subgroups (Opacity, Cardiomegaly and Effusion), which means that these populations have different distributions for these labels. Training on a balanced dataset or one that may have been sampled more Blackmale patients, for example, would have still induced generalizability biases on the under-represented population.

In our third analysis, we looked at the intersection between Black patients on private insurance and Black patients on Medicaid. Here, we notice the biggest potential biases occur when socioeconomic status is stacked on race. Looking simply at ‘no diagnosis’, we see that the AEquity for Black patients on Medicaid is 10.8, whereas the AEquity for Black patients that are privately insured is 9.2. This difference is both statistically significant (P < 0.05) and clinically significant because it implies there is a significant amount of complexity that occurs at baseline when comparing Black patients on Medicare to Black patients on private insurance. Second, we notice that the AEquity values are significantly higher for Black patients on Medicaid than Black patients that are privately insured in five of the class labels, but the opposite is only true for a single label, while the remaining labels are neutral. The intersection between race and socioeconomic status potentiates the largest potential for generalizability error, when compared to the intersectionality of race and sex or race and age.

Finally, we apply the relevant dataset intervention to Black patients on Medicaid with respect to the ‘No Finding’ or ‘None’ diagnosis. The AUC increases significantly, and we see that this change in the AUC increases by approximately 0.03 and is statistically significant (P < 1×10^-5^). To further dissect where this increase in AUC is coming from, we sought to further investigate the different fairness metrics including True Negative Rate, True Positive Rate, False Positive Rate, as well as further derived fairness metrics such as Precision, False Discovery Rate, and Predicted Prevalence.

We see that there is a clinically and statistically significant decrease in false positive rate for the diagnosis of “No Finding” (ΔFPR = 0.10) and a corresponding increase in true negative rate (ΔTNR = 0.10), with a clinically insignificant difference in false negative rate (ΔFNR = −0.01) and true positive rate (ΔTPR = −0.01). A lower false positive rate for the diagnosis of “No Finding” is equivalent to having a lower under-diagnosis bias rate. Furthermore, this translates into a higher precision, lower false discovery rate, and lower predicted prevalence. Changing the dataset gets at the root of the model’s problem described previously, which was under-diagnosis of patients in under-represented models. By taking the relevant mitigation measures, AEquity can reduce the false discovery rate and predicted prevalence of “No Finding”, thereby reducing the under-diagnosis bias in intersectional under-represented minorities.

### Analysis for Asian patients

In the MIMIC-CXR dataset, there are multiple races represented at lower sampling frequencies than White including Black and Asian. However, models trained on chest X-rays perform worse on Black patients compared to Asian patients. According to an analysis of AEquity values, this is because the sampling bias is compounded by complexity bias in the case of Black patients, whereas in Asian patients, the complexity bias may be less severe if present at all. When we calculate AEquity values with respect to Asian patients, we notice two things – first that the AEquity values for Asian patients tend to be lower than those of White patients. Second, the joint AEquity values between the Asian patients and White patients are also lower, indicating sampling bias. This is because the chest X-rays of Asian and White patients are also relatively similar compared to the difference between X-rays for Black and White patients. We have computed the AEquity for these patients below and reported them in Supplemental Figure 5.

## Supplementary Figures

**Fig. S1.**
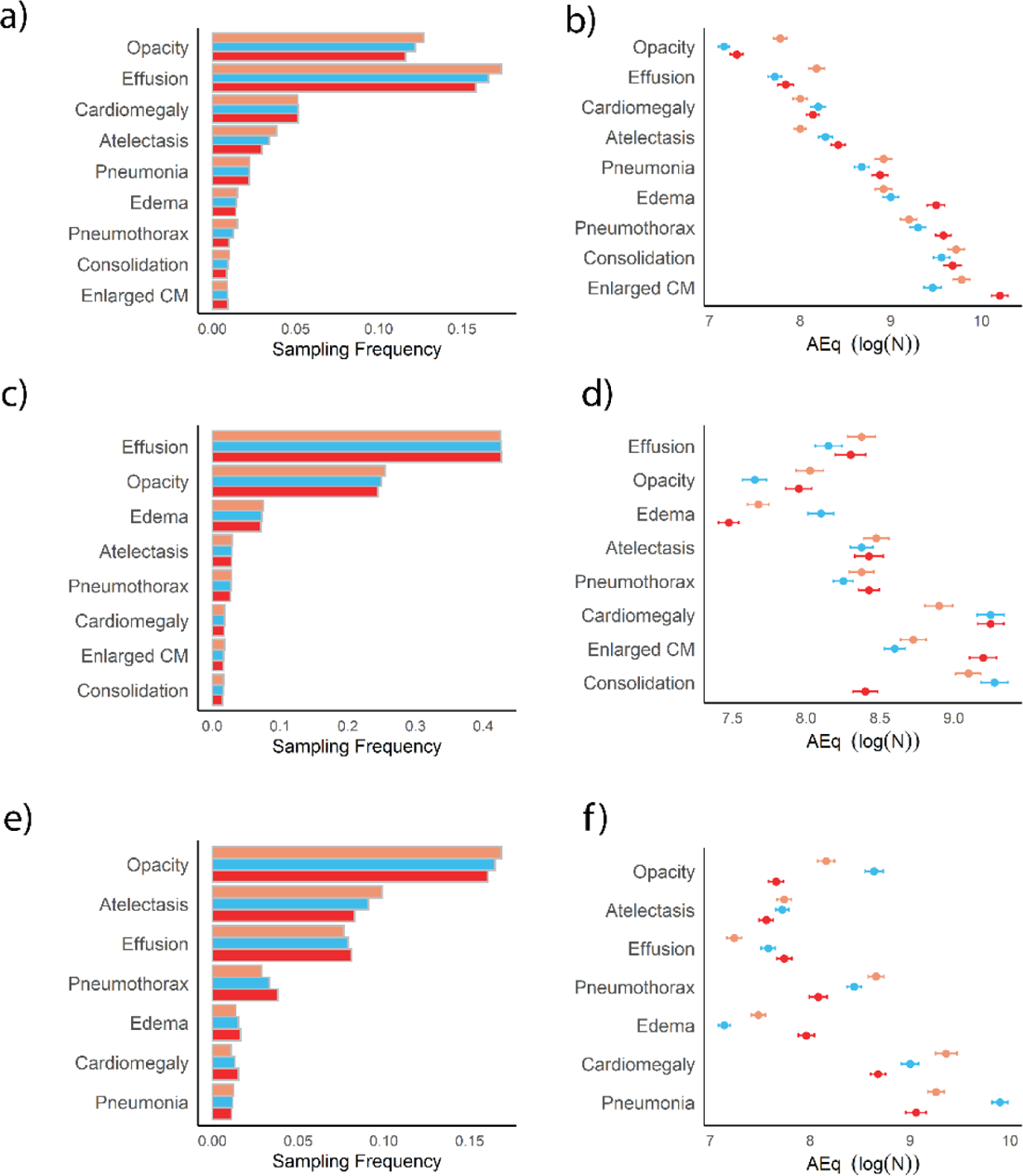
Replication on sex demographic. Red is female, orange is male and blue is the combined dataset. **a)** Histogram by class label of MIMIC-CXR and **b)** AEquity values for MIMIC-CXR. (R = −0.80, P = 4.94 x 10^-7^) **c)** Histogram by class label for CheXPert and **d)** AEquity values by class label for CheXPert. (R = −0.41, P = 8 x 10^-3^). **e)** Histogram by class label for ChestX-ray14, and **f)** Corresponding AEquity values for ChestX-ray14 (R = −0.41, P = 7 x 10^-3^).

**Fig S2.**
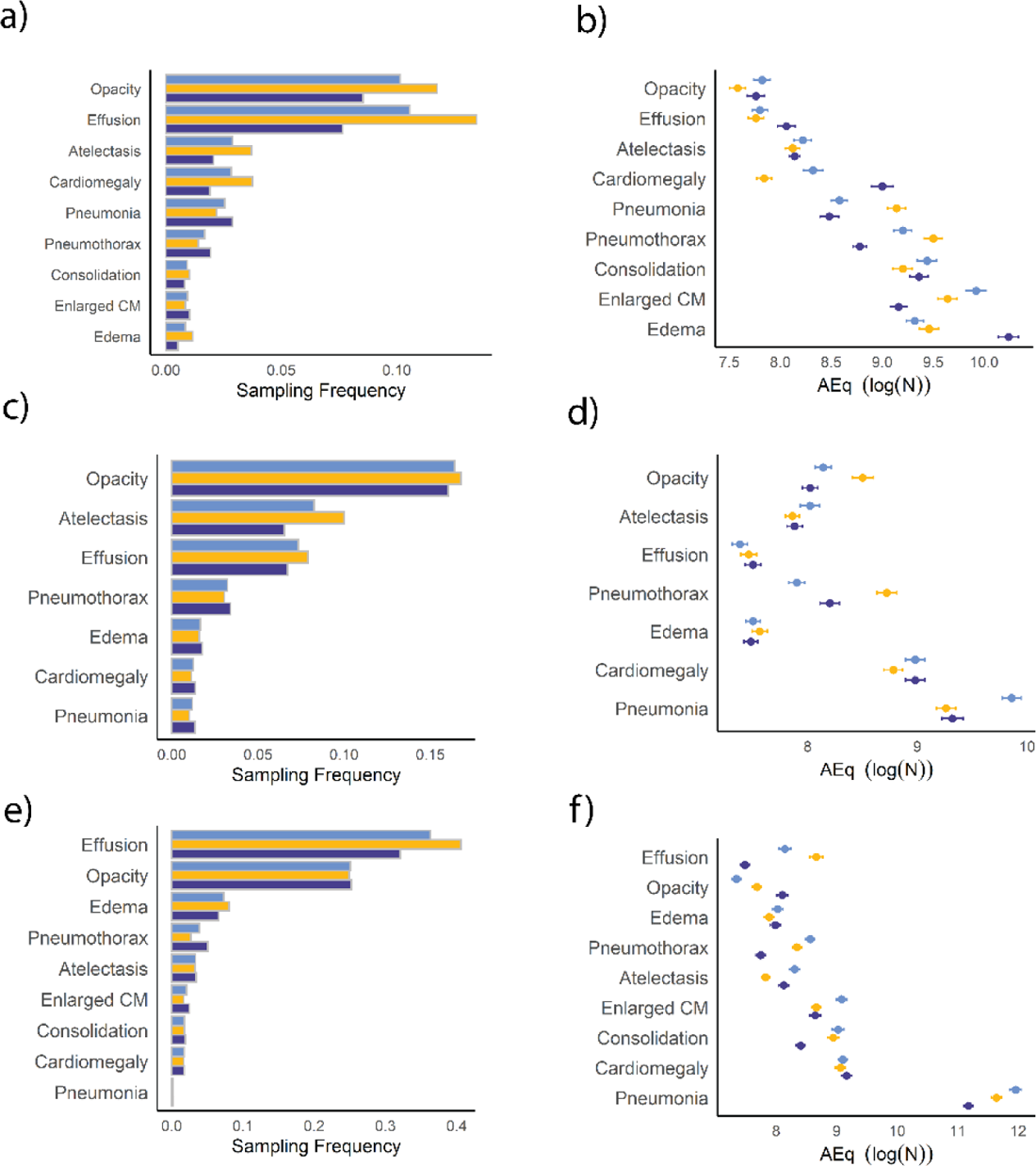
Replication on age demographic. Dark blue consists of individuals age (20-40) and yellow consists of individuals from 40-60. Light blue represents the joint dataset **a)** Histogram for individuals by age on MIMIC-CXR, **b)** Corresponding AEquity values for age on MIMIC-CXR (R =−0.88, P = 2 x 10^-8^). **c)** Histogram for individuals by age for ChestX-ray14 and **d)** corresponding AEquity values (R=−0.42, P = 3.4 x 10^-5^). **e)** Histogram for individuals by age on CheXPert and **f)** corresponding AEquity values (R = −0.32, P = 9.2 x 10^-3^).

**Fig S3.**
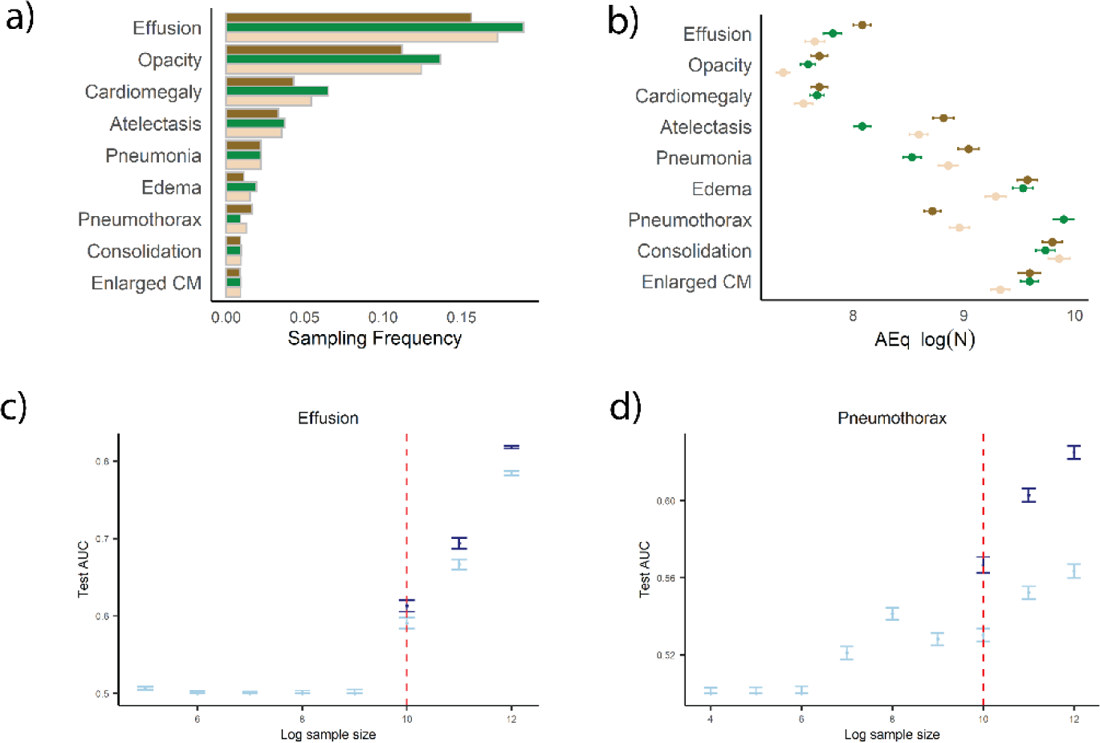
AEquity applied to the MIMIC-CXR dataset stratified by Insurance Status. Dark green represents privately insured, brown represents insured by Medicare, and beige represents joint data. **a)** Histogram of diagnostic frequencies by Insurance Status **b)** AEquity values by Insurance status. **c)** Intervention to increase dataset diversity improves performance in individuals on Medicare with effusion. **d)** Intervention to prioritize populations improves performance significantly in individuals on Medicare.

**Fig S4.**
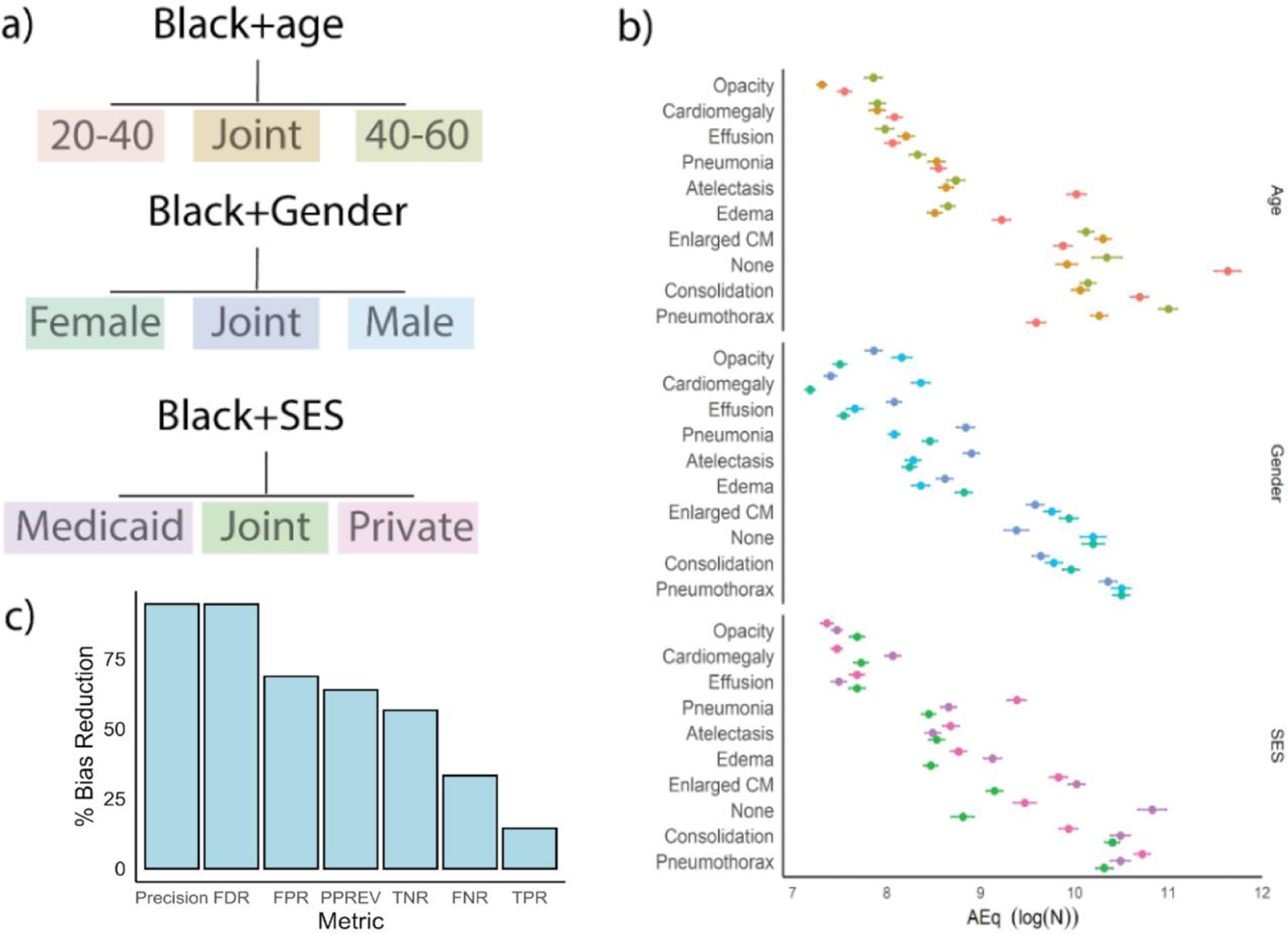
AEquity applied to intersectional patient populations. (a) Age – Black patients aged 20-40 are colored red, Black patients aged 40-60 are colored green, and the combined dataset is colored gold. Sex – Male Black patients are colored lighter turquoise, Female Black patients are colored light green, and the combined dataset is colored dark blue. Socioeconomic Status – Black patients on private insurance are colored pink, Black patients on Medicaid are colored dark purple, and the combined dataset is colored dark green. (b) AEq values calculated for each group across each label. (c) Interventions applied to Black patients on Medicaid and the resulting effects on different fairness metrics.

**Fig S5.**
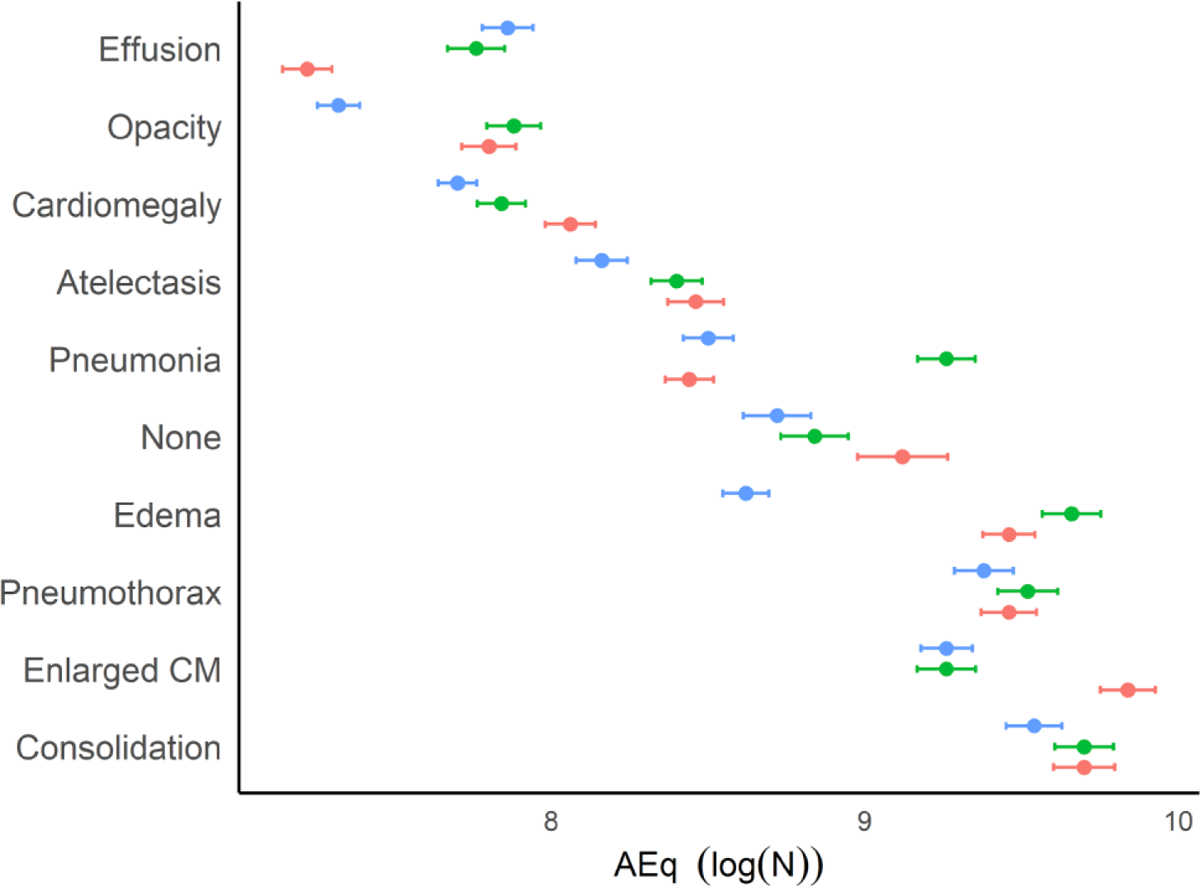
AEquity applied to patient population identifying as Asian. The bright red represents AEquity values calculated for individuals identifying as Asian. The green represents AEquity values calculated for individuals identifying as White, and the blue represents AEquity values calculated for a combined subset of individuals identifying as either Asian or White individuals. The AEquity of the combined subset is smaller than the individual subsets in eight of the 10 labels.

**Figure S6.**
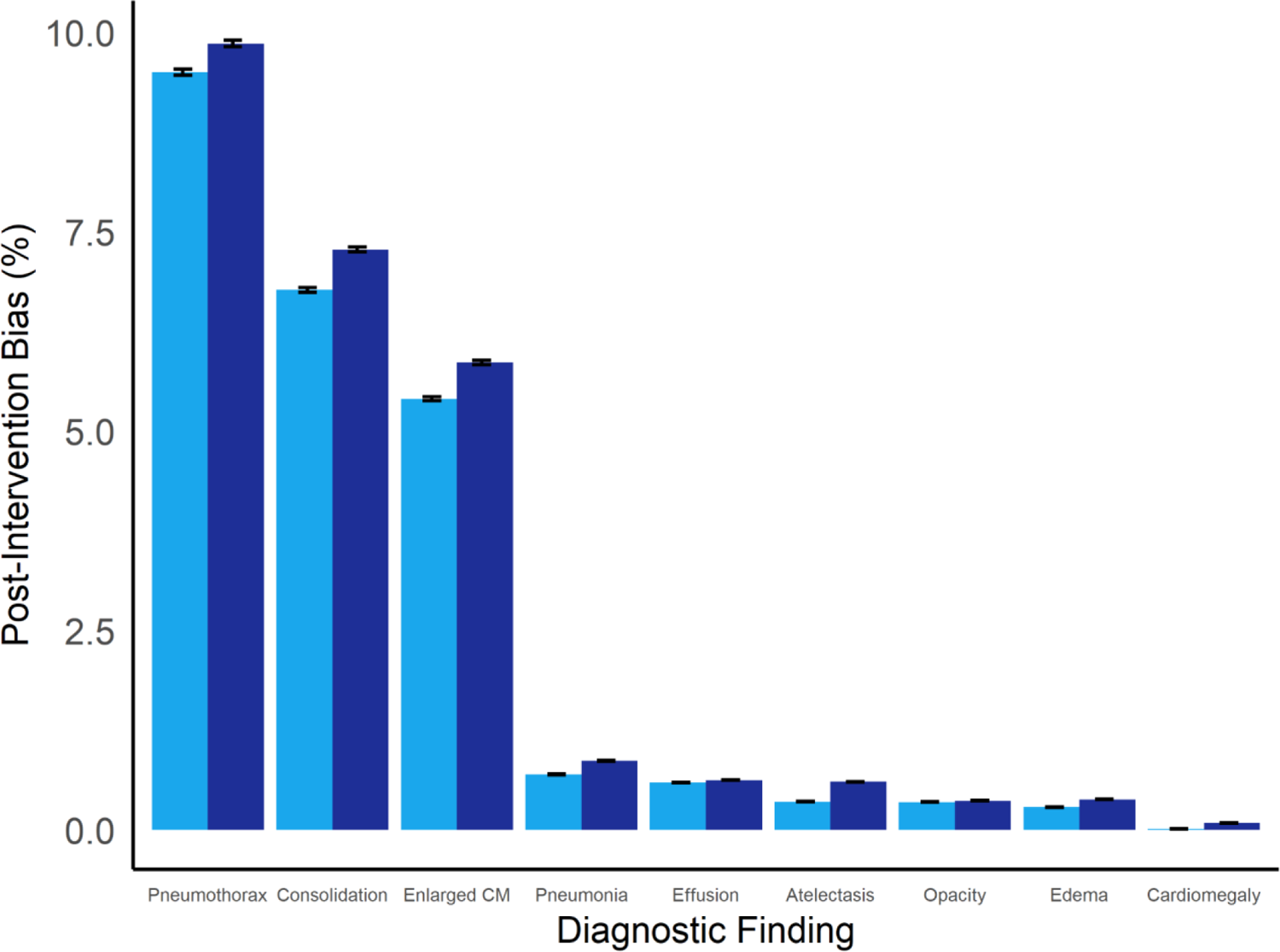
Measurement of bias by diagnosis in the radiograph dataset with VisionTransformers (86 M parameter VIT_B_16 with pre-training on IMAGENET) via area-under-the-curve. Light-blue is AEquity, Dark Blue is Balanced Empirical Risk Minimization.

**Figure S7.**
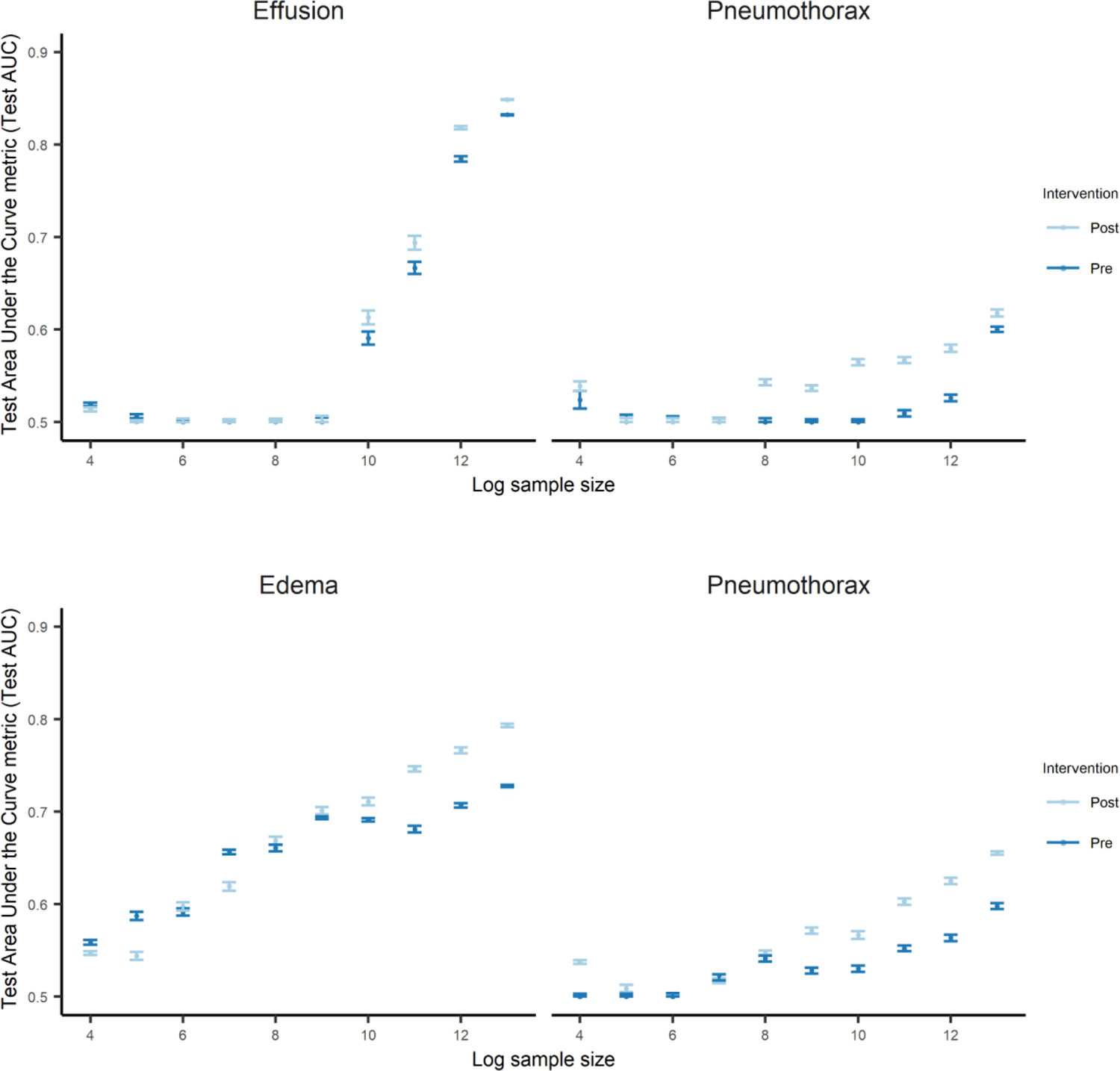
AEquity becomes increasingly precise as the number of samples increases and generalizes across different sample sizes when using small convolutional neural networks.

## Supplementary Tables

**Table S1.** Description of Datasets Utilized in Paper.

| Dataset | Description | Label/Outcome | Groups examined |
| --- | --- | --- | --- |
| MIMIC-CXR | 377,110 chest radiographs of 65,379 patients from the Beth Israel Deaconess Medical Center between 2011 and 2016. | Opacity, cardiomegaly, effusion, pneumonia, atelectasis, edema, enlarged cardiome-diastinum, consolidation, pneumothorax, no finding. | Race, sex, age, insurance status; Sex, age and insurance in Black patients. |
| Healthcare costs | All primary care patients ($N = 49,618$) enrolled in risk-based contracts from 2013 to 2015, from an unspecified large academic hospital. Data included demographics, visit information, costs, comorbidities and biomarkers. | Total costs, avoidable costs, comorbidity score. | Race. |
| ChestX-ray14 | 108,948 frontal chest radiographs from 32,717 patients from the NIH Clinical Center between 1992 and 2015. | Atelectasis, cardiomegaly, effusion, infiltration, mass, nodule, pneumonia, pneumothorax. | Sex, age. |
| CheXpert | 224,316 chest radiographs of 65,240 patients performed between October 2002 and July 2017 in both inpatient and outpatient centers affiliated with Stanford Hospital. | Opacity, atelectasis, effusion, pneumothorax, edema, cardiomegaly, pneumonia. | Sex, age. |
| NHANES | 47,261 samples (2000-2014) with demographic, laboratory, examination, and questionnaire features that could be automatically matched across different NHANES cycles. Follow-up mortality data is provided from the date of survey participation through December 31, 2015. | All-cause mortality for follow-up times of 1-year, 2-year, 3-year, 4-year, and 5-year. | Race. |

**Table S2.** Comparing AEquity to SOTA methods on MIMIC Chest X-ray classification tasks. We take a data-centric approach with simple training of a ResNet-50 pretrained model and achieve similar performance to state-of-the-art approaches that utilize a ResNet-50 backbone.

| Method | Test Area-Under-Curve |
| --- | --- |
| RadImageNet (2020)<sup>5</sup> | 0.71 |
| Wang et al. (2020)<sup>7</sup> | 0.74 |
| Ho et al. (2020)<sup>8</sup> | 0.74 |
| Yao et al (2018)<sup>9</sup> | 0.78 |
| Ours (RadImageNet) | 0.74 |
| Ours (Vit_B_16) | 0.83 |

**Table S3.**
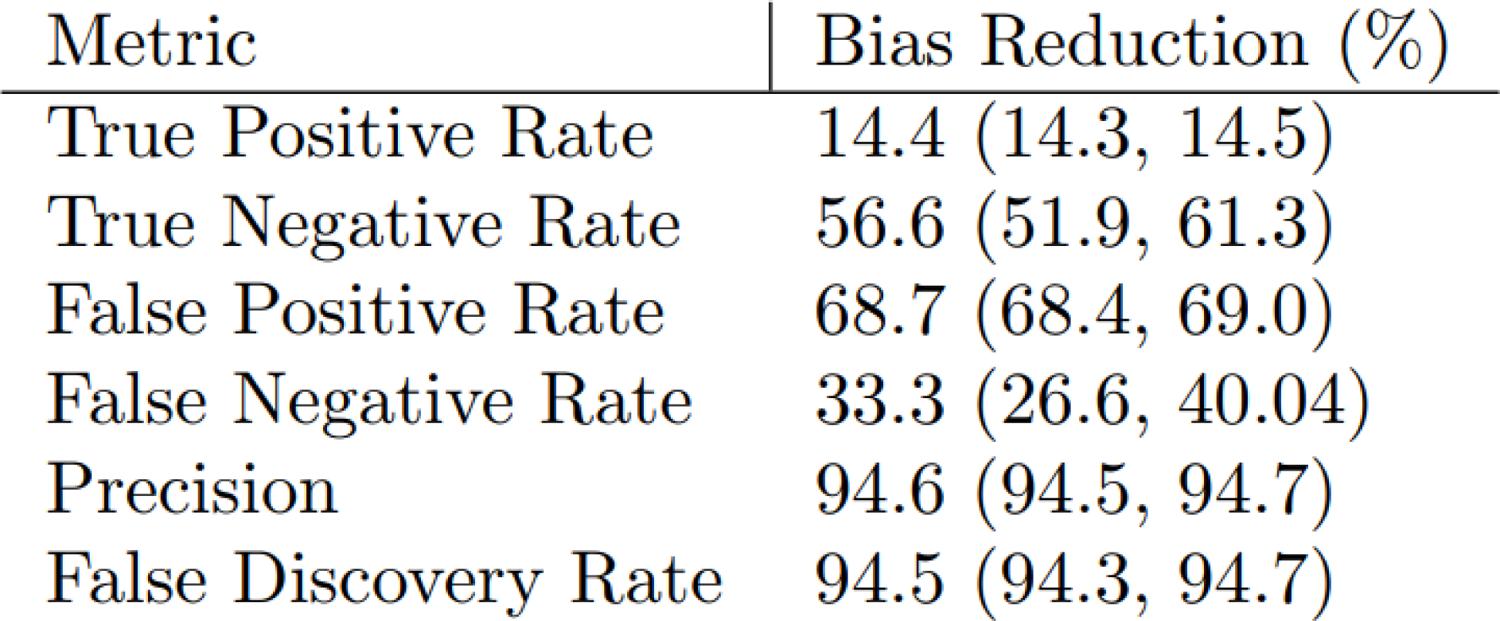
Measurement of bias and intervention effect by metric on an Intersectional Population We show the effect of the interventions on various metrics used to measure fairness for a patient population that is both Black and on Medicaid compared to White patients. With a dataset intervention, we have reduced the discrepancy between the two groups.

**Table S4.** AEquity metric by race and predicted risk level, for each outcome. We show the calculated AEquity metric for highest risk and lower risk individuals for Black and White races, for each of the three outcomes (total costs, avoidable costs and active chronic conditions).

| Metric | Category | Black | White | Difference |
| --- | --- | --- | --- | --- |
| Total Costs | high risk | 7.13 (7.05, 7.21) | 7.59 (7.47, 7.71) | 0.47 (0.36, 0.57) |
|  | low risk | 7.03 (6.97, 7.09) | 7.84 (7.72, 7.96) | 0.81 (0.73, 0.90) |
| Avoidable Costs | high risk | 7.09 (7.01, 7.17) | 7.50 (7.42, 7.58) | 0.41 (0.32, 0.49) |
|  | low risk | 7.91 (7.81, 8.01) | 7.09 (6.99, 7.19) | 0.81 (0.71, 0.92) |
| Active chronic conditions | high risk | 7.22 (7.12, 7.32) | 7.28 (7.18, 7.38) | 0.06 (-0.04, 0.16) |
|  | low risk | 6.97 (6.89, 7.05) | 7.31 (7.23, 7.39) | 0.34 (0.26, 0.43) |

**Table S5.**
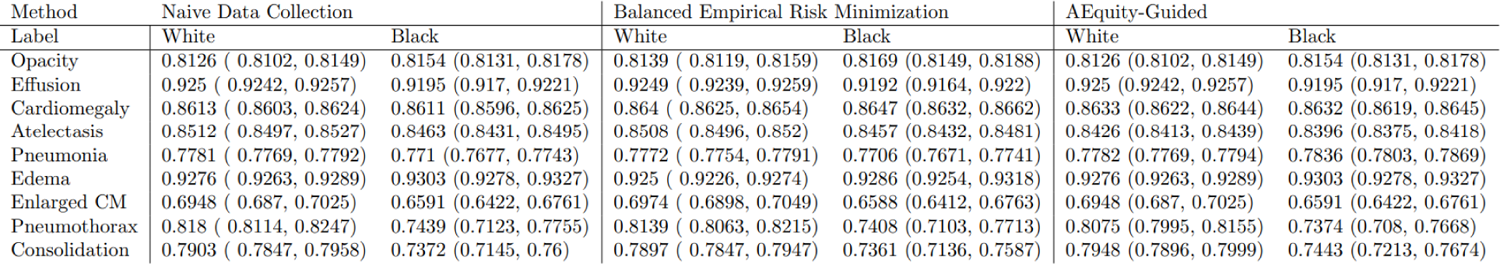
Measurement of bias and intervention effect by diagnosis in the radiograph dataset with VisionTransformers (86 M parameter VIT_B_16 with pre-training on IMAGENET)

